# Distinct Tau PET Dynamics in Early vs. Late Age-of-Onset Alzheimer’s disease

**DOI:** 10.64898/2026.01.25.26344738

**Authors:** Konstantinos Chiotis, Ganna Blazhenets, David N. Soleimani-Meigooni, Paul S. Aisen, Alinda Amuiri, Alireza Atri, Laurel Beckett, Michael Brickhouse, David G. Clark, Jeffrey L. Dage, Gregory S. Day, Ranjan Duara, Ani Eloyan, Tatiana Foroud, Neill R. Graff-Radford, Ian M. Grant, Dustin B. Hammers, Lawrence S. Honig, Erik C. B. Johnson, David T. Jones, Kala Kirby, Robert Koeppe, Joel H. Kramer, Walter A. Kukull, Julien Lagarde, Antoine Leuzy, Piyush Maiti, Joseph C. Masdeu, Mario F. Mendez, Erik Musiek, Kelly N. Nudelman, Chiadi U. Onyike, Meghan Riddle, Salma Rocha, Emily Rogalski, Stephen Salloway, Daniel R. Schonhaut, Sharon Sha, Ranjani Shankar, Alexander Taurone, Maryanne Thangarajah, Arthur W. Toga, Alexandra Touroutoglou, Raymond Scott Turner, Prashanthi Vemuri, Thomas S. Wingo, David A. Wolk, Kyle Womack, Jiaxiuxiu Zhang, Maria C. Carrillo, Bradford C. Dickerson, Liana G. Apostolova, Renaud La Joie, Gil D. Rabinovici, the Longitudinal Early-onset Alzheimer’s Disease Study consortium and the Alzheimer’s Disease Neuroimaging Initiative

## Abstract

Early-onset Alzheimer’s disease (EOAD) and Late-onset AD (LOAD) differ in clinical presentations and rates of progression. We aimed to compare baseline and longitudinal tau PET burden, and their relationship with clinical variables in amyloid-PET positive, cognitively impaired participants from the Longitudinal Early-Onset Alzheimer’s Disease Study (EOAD; n=390) and Alzheimer’s Disease Neuroimaging Initiative (LOAD; n=211). Patients with EOAD showed higher baseline tau PET retention, broader neuroanatomical involvement and faster accumulation rates over time compared to LOAD, after adjusting for amyloid load and clinical stage. Tau PET showed stronger correlations with baseline amyloid burden and clinical measures of global cognition and function in EOAD than LOAD. We conclude that earlier age of onset in AD is linked to a more aggressive tauopathy, which in turn is a primary driver of clinical decline. These findings suggest that optimal therapeutic targets and strategies may differ between EOAD and LOAD.

**One Sentence Summary:** Younger patients with Alzheimer’s disease show more aggressive tau spread, suggesting age of onset defines distinct disease pathways with key clinical implications.

## Introduction

Alzheimer’s disease (AD), the leading cause of dementia, presents a significant global public health challenge as the most prevalent neurodegenerative disorder and a major contributor to disability and dependence among older adults worldwide (*1*). AD is classified by symptom onset age as early-(EOAD, symptom onset age<65) or late-onset (LOAD, symptom onset age>65). EOAD accounts for about 5% of all cases, (*2*) often affecting individuals in their prime working years, and is frequently associated with atypical (i.e., non-amnestic) clinical presentations, delayed diagnosis, and faster cognitive decline compared to LOAD (*3*). Importantly, only ∼5% of patients with EOAD carry known pathogenic mutations associated with familial AD (*4*), whereas most present with sporadic disease at a young age due to unknown genetic or environmental risk factors.

Although the 65-year cutoff to differentiate EOAD from LOAD is largely arbitrary, evidence suggests that early and late-onset presentations of AD diverge in meaningful ways (*5*). Both EOAD and LOAD feature pathological accumulation of amyloid-beta (Aβ) and tau proteins in the brain, forming extracellular plaques and intraneuronal neurofibrillary tangles, respectively. However, neuropathological studies show EOAD typically exhibits more extensive Aβ and tau aggregation compared to LOAD (*6*), particularly in neocortical regions, implying distinct biological factors underlying differences in clinical phenotypes. Conversely, patients with EOAD show fewer age-associated co-pathologies (e.g., TDP-43 pathology, argyrophilic grain disease, hippocampal sclerosis, or cerebrovascular lesions) than older AD patients (*7*), suggesting that a more “pure” form of AD may drive cognitive decline. A better understanding of the biomarker profiles and molecular mechanisms distinguishing EOAD and LOAD is particularly relevant in the emergent era of disease-modifying therapies, recognizing that optimal therapeutic strategies may differ in patients with EOAD and LOAD.

The introduction of tau-specific PET tracers substantially advanced our understanding of AD’s spatiotemporal progression, particularly in LOAD (*8*). Yet, the differences in tau PET load between EOAD and LOAD remain underexplored. Initial findings from modest-sized, cross-sectional studies indicate greater tau burden in EOAD than LOAD (*9–14*). However, these results may be confounded by differences in disease stage at diagnosis, complicating the interpretation of whether discrepancies in tau pathology reflect more advanced disease stages in EOAD (*15*) or inherent biological differences associated with age of symptom onset. Supporting the latter, recent in vitro studies suggest that tau species linked to earlier age of onset exhibit more aggressive spreading patterns (*16*), potentially explaining the higher tau load observed in EOAD.

This study utilizes tau PET imaging data from two of the largest longitudinal AD cohorts—the Longitudinal Early-onset Alzheimer’s Disease Study (LEADS), which focuses on sporadic EOAD, and the Alzheimer’s Disease Neuroimaging Initiative (ADNI), primarily comprising sporadic LOAD participants—to investigate the differential tau burden between EOAD and LOAD. By integrating longitudinal multimodal data, this research aims to delineate shared and distinct features of tau pathology, taking into account the continuous effects of age alongside differences in disease stage and Aβ load between age groups. This comprehensive approach enables modeling of tau accumulation trajectories, providing a unique opportunity to compare tau progression rates between EOAD and LOAD.

## Results

### Clinical characteristics

Table 1 presents the demographic and clinical characteristics of EOAD and LOAD participants. We identified 390 participants with EOAD from the LEADS cohort and 211 with LOAD from the ADNI cohort. In the EOAD group, 20% presented with a predominantly non-amnestic syndrome, whereas cases with LOAD almost exclusively presented with an amnestic syndrome. Both groups were quantitatively Aβ PET positive (>24.4 Centiloids [CL]) and diagnosed with Mild Cognitive Impairment (MCI) or dementia at baseline. The EOAD group had a higher proportion of females (54% vs 42%, p=0.006). A significantly higher proportion of participants with EOAD were diagnosed at baseline with dementia compared to those with LOAD (72% vs 36%, p<0.001). At baseline, the EOAD group exhibited higher Aβ PET tracer binding (97.9±27.1 CL vs 85.3±33.5 CL, p<0.001) and more severe impairment across global cognitive measures (i.e., Mini-Mental State Examination [MMSE] and Clinical Dementia Rating sum-of-boxes [CDR-SB]) than the LOAD group. Longitudinally, tau PET scans were available for 245 participants with EOAD and 99 with LOAD. The follow-up interval for the LEADS participants was shorter than that for ADNI (2.1±1.0 vs 2.8±1.7, p<0.001), although the number of follow-up time points did not differ significantly between groups.

**Table 1.** Clinical characteristics of the patient sample.

|  | **EOAD (LEADS), N = 390<sup>1</sup>** | **LOAD (ADNI), N = 211<sup>1</sup>** | **p-value<sup>2</sup>** |
| --- | --- | --- | --- |
| **Baseline characteristics** |  |  |  |
| **Age in years at baseline** | 59.16 ± 4.03 | 77.65 ± 6.59 | **<0.001** |
| **Sex** |  |  | **0.006** |
| Male | 178 (45.64%) | 122 (57.82%) |  |
| Female | 212 (54.36%) | 89 (42.18%) |  |
| **Education in years** | 15.64 ± 2.45 | 16.03 ± 2.60 | 0.069 |
| **APOE-ε4 allele** |  |  | 0.081 |
| Carrier (≥ 1 allele) | 207 (55.20%) | 122 (63.21%) |  |
| Non carrier | 168 (44.80%) | 71 (36.79%) |  |
| Missing | 15 | 18 |  |
| **Race/Ethnicity** |  |  | 0.140 |
| White, not Hispanic or Latino | 340 (87.63%) | 165 (82.50%) |  |
| White, Hispanic or Latino | 10 (2.58%) | 9 (4.50%) |  |
| Black or African American | 22 (5.67%) | 19 (9.50%) |  |
| Asian | 10 (2.58%) | 2 (1.00%) |  |
| More than one | 6 (1.55%) | 5 (2.50%) |  |
| Missing | 2 | 11 |  |
| **Clinical stage at baseline** |  |  | **<0.001** |
| Dementia | 282 (72.31%) | 77 (36.49%) |  |
| MCI | 108 (27.69%) | 134 (63.51%) |  |
| **Clinical syndrome at baseline** |  |  | **<0.001** |
| Amnestic | 313 (80.26%) | 133 (99.25%) |  |
| Non-amnestic (including<br>logopenic variant of primary<br>progressive aphasia or posterior<br>cortical atrophy) | 77 (19.74%) | 1 (0.75%) |  |
| Missing | 0 | 77<sup>‡</sup> |  |
| **Centiloids at baseline** | 97.88 ± 27.14 | 85.32 ± 33.51 | **<0.001** |
| **CDR-SB at baseline** | 3.87 ± 1.99 | 2.77 ± 2.38 | **<0.001** |
| Missing | 3 | 0 |  |
| **CDR Global at baseline** |  |  | **<0.001** |
| 0 | 5 (1.29%) | 10 (4.74%) |  |
| 0.5 | 228 (58.91%) | 156 (73.93%) |  |
| 1 | 152 (39.28%) | 41 (19.43%) |  |
| > 1 | 2 (0.52%) | 4 (1.89%) |  |
| Missing | 3 | 0 |  |
| **MMSE** | 21.34 ± 5.43 | 25.63 ± 3.68 | **<0.001** |
| Missing | 7 | 0 |  |
| **<u>Longitudinal characteristics</u>** |  |  |  |
| **Participants with follow-up imaging** | 245 | 99 |  |
| **Follow-up interval in years** | 2.06 ± 1.00 | 2.80 ± 1.71 | **<0.001** |
| **Number of longitudinal scans** | 2.63 ± 0.79 | 2.71 ± 0.82 | 0.400 |

<sup>1</sup>Mean ± SD; n (%); <sup>2</sup>Welch Two Sample t-test; Pearson's Chi-squared test. <sup>‡</sup> The clinical syndrome for dementia cases in ADNI is not documented and cases with atypical variants of AD are not the focus of the ADNI study. CDR = Clinical Dementia Rating; MCI = Mild Cognitive Impairment; MMSE = Mini Mental State Examination

We also identified cognitively normal individuals with Aβ-negative (n=85 from LEADS and n=328 from ADNI) and positive scans (n=9 from LEADS and n=125 from ADNI). These data were used for comparisons of tau PET binding patterns with those observed in the EOAD and LOAD groups. A detailed comparison of their clinical characteristics is in Supplementary Table 1.

### Baseline tau PET levels

We assessed cross-sectional differences in baseline ^18^F-Flortaucipir tau PET binding between EOAD and LOAD groups. Binding levels were expressed as SUVR in CenTauR target regions of interest (ROIs) relative to the inferior cerebellar CenTauR ROI, and also linearly transformed to CenTauR Z scores or CenTauR (CTR) units using the Joint Propagation Model approach, as previously detailed (*17, 18*). EOAD participants exhibited significantly higher binding across all ROIs, regardless of MCI or dementia stage (Fig. 1A-D, Supplementary Fig. 1). This was less evident in the mesial temporal lobe, where there was substantial overlap between age groups. A higher proportion of patients with EOAD surpassed the tau PET positivity threshold of 2 CenTauR Z-scores in all ROIs compared with those with LOAD. Both age groups showed substantially greater proportions of patients above this threshold in all ROIs, compared to cognitively normal groups. Binding in the LEADS Aβ-negative cognitively normal group was significantly lower compared to the ADNI group in the mesial temporal, meta-temporal, and universal ROIs. No significant differences were observed between Aβ-positive cognitively normal groups in the two cohorts. Notably, the small size of the LEADS Aβ-positive cognitively normal group (n=9) limits robust statistical comparisons (Fig. 1, Supplementary Table 1).

**Fig 1.**
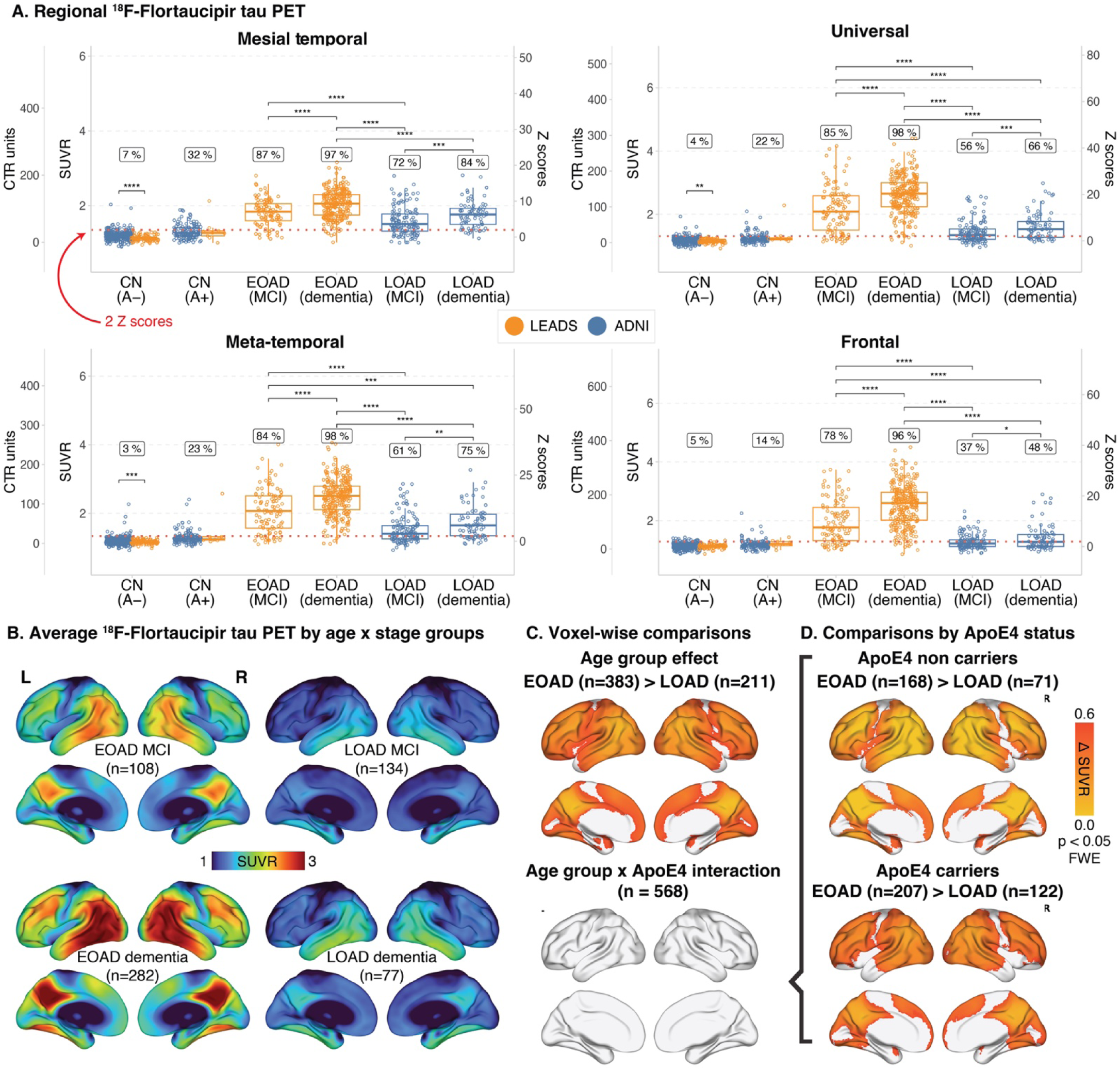
Group comparisons of baseline ^18^F-Flortaucipir binding across EOAD and LOAD patient groups in the standard CenTauR ROIs. (A) and voxel-wise analyses in the whole sample (B, C) and within the *APOE*-ε4 subgroups (D). In the ROI-based comparisons, formal statistical tests were restricted to the EOAD and LOAD subgroups, as well as cognitively normal (CN) participants, stratified by Aβ PET status using a threshold of 24.4 CL to define Aβ positive (A+) and negative (A-) subgroups. The percentages above each boxplot indicate the proportion of individuals exceeding the 2 CenTauR Z-score threshold for binding load, based on Villemagne et al. (*17*). The binding data are also presented in CenTauR (CTR) units, following the Joint Propagation Model approach by Leuzy et al. (*18*). In the voxel-wise comparisons (C, D), patients missing baseline MMSE scores were excluded. These voxel-wise analyses were adjusted for baseline Centiloid, MMSE, sex, and years of education, and corrected for multiple comparisons using the FWE rate approach, as implemented in SPM12 software.

Voxel-wise average tau PET maps for MCI and dementia stages in EOAD and LOAD highlighted significant, large effect size differences in ^18^F-Flortaucipir binding between groups in widespread areas except the mesial temporal lobe and primary cortices (Fig. 1B). These findings were consistent in patients with global CDR scores of 0.5 or 1 (Supplementary Fig. 2). Voxel-wise linear regression, adjusting for sex, years of education, baseline MMSE, and baseline Aβ PET burden, showed significantly higher tau binding in EOAD than LOAD, particularly in widespread frontal, temporal, and parietal areas (Fig. 1D). The primary cortices and mesial temporal lobes were exceptions to this pattern.

Conversely, LOAD showed higher ^18^F-Flortaucipir PET binding in the basal ganglia and choroid plexus (Supplementary Fig. 3)–regions associated with “off-target” (i.e., non-tau related) ^18^F-Flortaucipir binding and known to show age-related increases in cognitively unimpaired controls (*19*).

No significant age group by *apolipoprotein E* ε4 (*APOE*-ε4) interaction effects were found (Fig. 1C). Voxel-wise comparisons stratified by *APOE*-ε4 status showed similar neocortical regions with higher binding in EOAD than LOAD (Fig. 1D), consistent with the whole sample.

### Association of age as a continuous measure with tau

Across both EOAD and LOAD, age was significantly negatively associated with ^18^F-Flortaucipir tau PET binding load in the mesial temporal and universal ROIs (Fig. 2A), while adjusting for sex, years of education, baseline MMSE, and baseline Aβ PET levels. Associations within MCI and dementia subgroups of the two cohorts are shown in Supplementary Fig. 4. In the whole sample (EOAD and LOAD), and after adjusting for the above-mentioned covariates, age was negatively associated with ^18^F-Flortaucipir binding in widespread cortical areas, while a significant positive association was found in the basal ganglia and choroid plexus, consistent with the group-level comparisons (Supplementary Fig. 5).

**Fig. 2.**
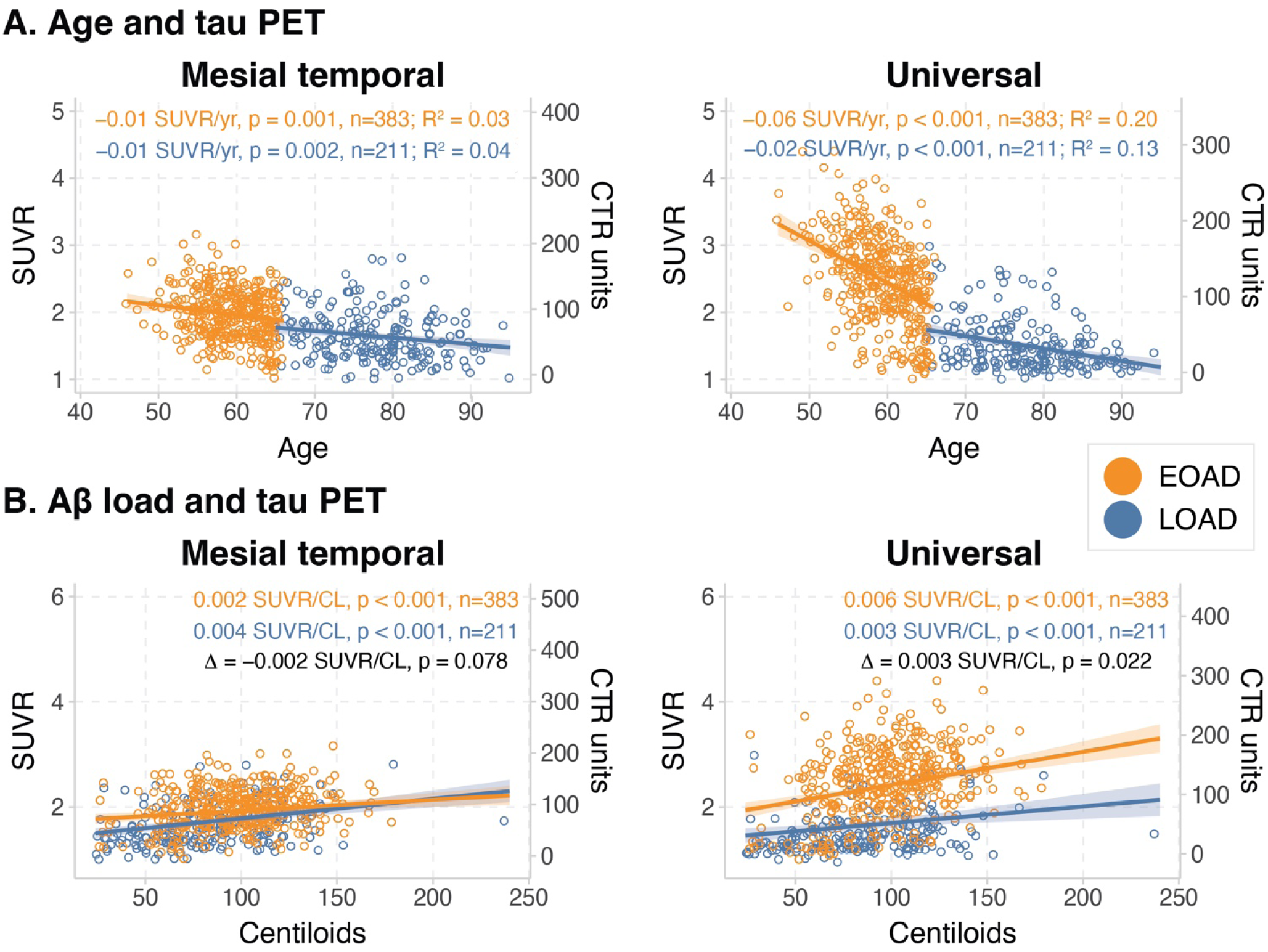
Scatterplots showing the association between age. (A) and Aβ PET binding levels (B) lobe and universal masks across the two age groups (i.e., EOAD and LOAD). The binding data are also presented in CenTauR (CTR) units, following the Joint Propagation Model approach by Leuzy et al. (*18*). Linear models were adjusted for MMSE, sex, and years of education, with age models additionally including baseline Centiloid as a covariate. Patients missing baseline MMSE scores were excluded. The linear coefficients for EOAD and LOAD are shown in orange and blue, respectively, and for the interaction in black. The was calculated using the partial coefficient of determination (R²) from the multivariate linear model.

### Association of Aβ PET levels with tau

Next, we assessed associations of Aβ and ^18^F-Flortaucipir tau PET binding in EOAD and LOAD. Although both groups showed positive associations, a significant interaction between Aβ PET load and age group was observed in predicting tau PET load in the universal ROI but not in the mesial temporal ROI, while adjusting for baseline MMSE, sex, and years of education. Specifically, EOAD showed a steeper association of Aβ with tau PET levels, than LOAD.

### Association of cognitive scores with tau levels

We further explored the relationship between cognitive performance—measured by MMSE and CDR-SB—and ^18^F-Flortaucipir tau PET binding within EOAD and LOAD. Greater cognitive impairment was associated with higher tau loads across the CenTauR ROIs in both groups (Fig. 3A, B). A significant interaction emerged between cognitive score and age group in predicting ^18^F-Flortaucipir binding, adjusting for baseline Aβ PET levels, sex, and years of education. Across different levels of impairment, EOAD showed higher ^18^F-Flortaucipir binding than LOAD group in all ROIs except the mesial temporal lobe. In voxel-wise models, this interaction was significant for CDR-SB but not MMSE, particularly in frontal areas (Fig. 3A, B). Separate voxel-wise models demonstrated distinct patterns: in LOAD, significant associations were localized to temporal with or without parietal areas, whereas in EOAD, they were more widespread across neocortical regions.

**Fig. 3.**
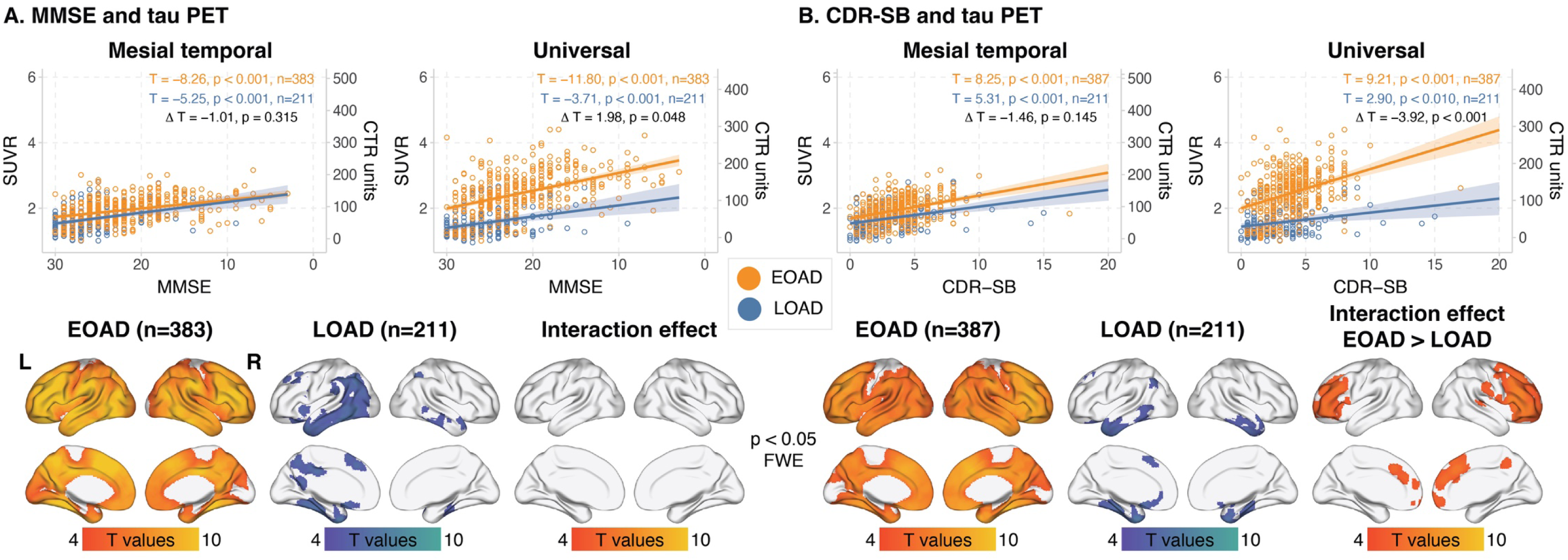
Association between baseline MMSE score. (A) and CDR-SB (B) with 18F-Flortaucipir tau PET binding in the CenTauR mesial temporal and universal ROIs and in voxel-based analyses. The binding data are also presented in CenTauR (CTR) units, following the Joint Propagation Model approach by Leuzy et al. (*18*) Patients missing baseline MMSE or CDR-SB scores were excluded from the respective analyses. The linear coefficients for EOAD and LOAD are shown in orange and blue, respectively, and for the interaction in black. The voxel-wise analyses were adjusted for baseline Centiloid, sex, and years of education, and corrected for multiple comparisons using the FWE rate approach, as implemented in SPM12 software.

### Longitudinal difference in tau accumulation rates

We employed linear mixed-effects models to examine the interaction between time from baseline and age group (EOAD vs. LOAD) in predicting longitudinal changes in ^18^F-Flortaucipir tau PET binding. These models adjusted for participant-specific changes, baseline Aβ PET load, MMSE, sex and years of education across the CenTauR ROIs, and revealed a significant interaction (Fig. 4A, Fig. 5A, Supplementary Table 2 and Supplementary Fig. 6). Simple slope analyses indicated that both age groups exhibited significant increases in ^18^F-Flortaucipir binding across all ROIs over time, except for the frontal ROI, where LOAD showed no significant change. In EOAD, the rates ranged from 0.06±0.00 SUVR/year in the mesial temporal ROI to 0.10±0.01 SUVR/year in the frontal ROI. For LOAD, rates were 0.02±0.01 SUVR/year in both the mesial temporal and frontal ROIs. In the universal ROI, rates were 0.07±0.01 SUVR/year for EOAD and 0.03±0.01 SUVR/year for LOAD. The rates of increase were significantly steeper in EOAD in all ROIs, and the interaction effect remained significant after adjusting for baseline ^18^F-Flortaucipir levels. This pattern persisted within the MCI and dementia subgroups, although in the relatively small LOAD dementia subgroup, statistically significant increases in ^18^F-Flortaucipir binding were not detected.

**Fig. 4.**
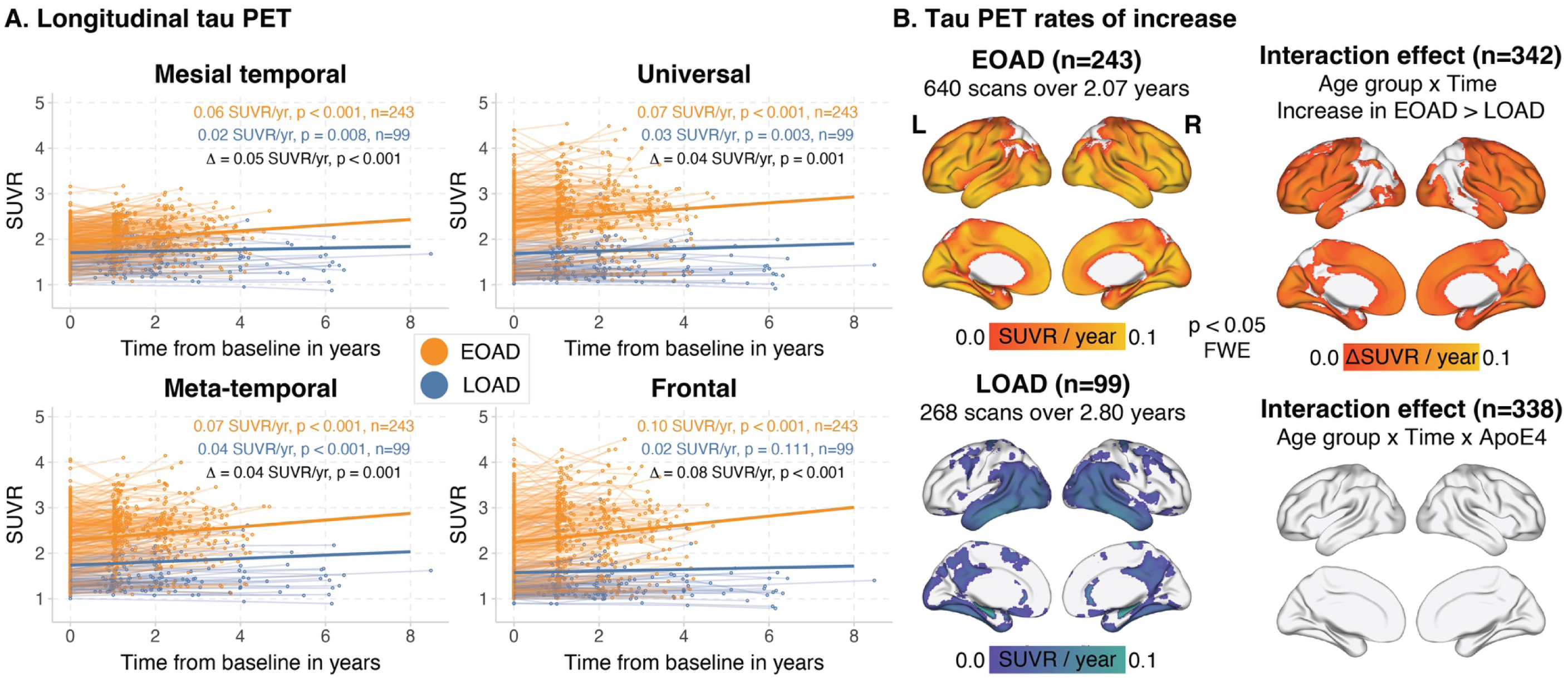
Longitudinal changes in ^18^F-Flortaucipir tau PET binding over the follow-up interval in patients with EOAD and LOAD depicted using spaghetti plots for CenTauR ROIs. (A) and voxel-wise analyses (B). The ROI-based linear mixed-effects model coefficients for EOAD and LOAD are shown in orange and blue, respectively, and for the interaction in black. Models were adjusted for baseline MMSE score, Centiloid, sex, and years of education, with random intercepts and slopes at the individual patient level. Patients missing baseline MMSE scores were excluded from the statistical modeling. The voxel-wise analyses were corrected for multiple comparisons using the FWE rate at the peak level, as implemented in Voxelstats software (*20*).

**Fig. 5.**
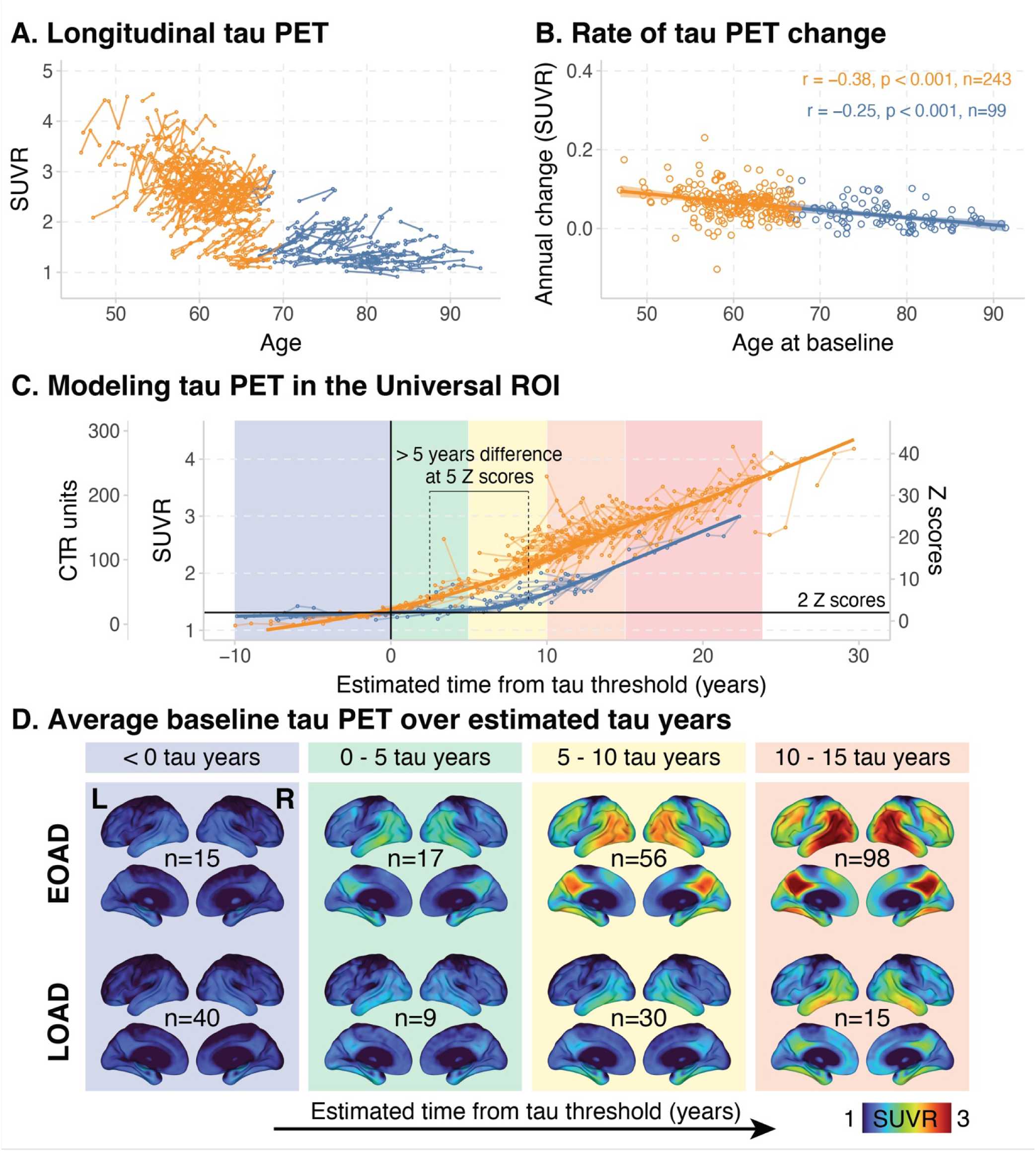
Longitudinal modeling of ^18^F-Flortaucipir tau PET binding in the Universal ROI in EOAD and LOAD, illustrated by trajectory plots for individual patients. (A), scatterplots showing age at baseline relative to the estimated annual rate of change in tau PET binding from the linear mixed-effects models (B), modeled group trajectories as a function of tau duration from the CenTauR threshold of 2 Z-scores using the SILA pipeline in the Universal ROI (C), and average baseline ^18^F-Flortaucipir tau PET scans across tau duration groups (D). The binding data are also presented in CenTauR (CTR) units, following the Joint Propagation Model approach by Leuzy et al. (*18*) The linear mixed-effects models (B) were adjusted for baseline MMSE score, Centiloid, sex, and years of education, with random intercepts and slopes at the individual patient level. Patients missing baseline MMSE scores were excluded from the statistical modeling. The estimated annual rates of change in ^18^F Flortaucipir tau PET binding were derived from the sum of the fixed and random coefficients for each individual. The Pearson correlation coefficients for EOAD and LOAD are shown in orange and blue, respectively. The SILA models were anchored to the exploratory 2 CenTauR Z-score threshold, which was used to define ^18^F-Flortaucipir tau PET binding positivity, and the trajectories are modeled relative to time in years from crossing the threshold.

Voxel-wise analyses with a similar design revealed a significant interaction (Fig. 4B), predominantly in cortical ROIs while largely sparing the temporoparietal lobes. A steeper increase was detected in ^18^F-Flortaucipir binding in EOAD. In separate models for each age group, LOAD showed significant binding increases primarily within the temporoparietal cortices, whereas EOAD exhibited increases in widespread cortical areas, sparing the lateral parietal lobes.

The three-way interaction between age group, time from baseline, and *APOE*-ε4 carrier status showed no significant effect (Fig. 4B). Stratified analyses by *APOE*-ε4 carrier status showed similar patterns of ^18^F-Flortaucipir binding increases as in the whole sample (Supplementary Fig. 7). *APOE*-ε4 carriers showed more widespread binding increases in both age groups. The smallest LOAD non-carrier group (n=32) showed limited increases over time.

Separate linear mixed-effects models with the above-mentioned design were employed for EOAD and LOAD to estimate the annual rates of change for each participant. The rates of tau change were significantly inversely associated with age at baseline in both cohorts, with similar slopes across most regions (Fig. 5B and Supplementary Fig. 6). No association was observed in LOAD in the frontal ROI, consistent with the lack of significant increases in binding in that ROI.

We applied the sampled iterative local approximation (SILA) algorithm (*21*) to model longitudinal ^18^F-Flortaucipir tau PET trajectories in EOAD and LOAD. Tau positivity duration curves were anchored at zero, defined by the tau positivity threshold in the universal ROI (i.e., 2 CenTauR Z-scores). We observed that EOAD had a steeper increase, indicating faster increases in ^18^F-Flortaucipir binding once the positivity threshold was reached. In contrast, patients with LOAD exhibited more than five years’ delay before reaching similar levels. Averaging baseline ^18^F-Flortaucipir scans across tau duration groups revealed widespread neocortical involvement in EOAD soon after crossing the threshold. In contrast, even in the most advanced LOAD, the tracer binding in extra-temporoparietal regions remained comparatively low. Similar patterns of accelerated increases in ^18^F-Flortaucipir binding in EOAD compared to LOAD were observed across other CenTauR ROIs (Supplementary Fig. 8).

## Discussion

The heterogeneity observed in AD, characterized by variations in clinical presentation, rates of cognitive decline, and neuropathological burden, is significantly influenced by age at onset (*5,6, 22, 23*). Despite this variability, few studies have thoroughly compared tau propagation dynamics in participants with EOAD and LOAD. This study addresses this gap by examining the temporal progression and spatial distribution of tau pathology in EOAD versus LOAD, using longitudinal tau PET data from two large, multi-site observational studies: LEADS and ADNI.

Our findings revealed that patients with EOAD exhibit not only higher tau levels, consistent with previous studies (*9, 10, 12–14*), but also faster tau accumulation. The divergence in tau trajectories between EOAD and LOAD parallels the higher Aβ burden in EOAD (*24*), suggesting the possibility of common upstream mechanisms influencing both pathologies. These results align with earlier neuropathology studies showing greater pathology burden in EOAD (*6, 23*). However, the magnitude of Aβ differences was smaller than that for tau. Our study advances the existing evidence by directly comparing tau load while adjusting for Aβ differences in large samples of patients studied *in vivo,* at different symptomatic stages. The significantly greater tau levels in EOAD across Aβ levels underscore tau load as the dominant neuropathological factor distinguishing the two age groups. This suggests a more aggressive tauopathy in younger patients with AD, likely driving the faster neurodegeneration observed (*25*), given the established relationship between tau buildup and neuronal loss (*26, 27*).

A strong body of evidence shows that patients with EOAD typically present at more severe stages of cognitive impairment than LOAD, often due to delayed diagnosis (*3, 28*). This delay likely stems from lower clinical suspicion given the younger age and atypical clinical phenotypes of EOAD, which often lack the episodic memory deficits common in LOAD (*3, 29*). Several studies comparing EOAD and LOAD consistently report more advanced disease stages in EOAD at diagnosis, complicating fair comparisons of neuropathological burden in modest-sized biomarker studies (*10*). Our study, based on two of the largest neuroimaging cohorts in EOAD and LOAD, overcomes this by adjusting for cognitive impairment and enabling detailed subgroup analyses. Our findings highlight that EOAD’s greater tau pathology is not solely due to clinical stage differences but rather reflects inherent age-related tau propagation differences. These differences extend beyond tau load to spatial distribution of the pathology. Specifically, EOAD shows broader neocortical tau as early as the MCI stage, whereas in LOAD, tau pathology remains more centered in temporoparietal regions, even at the dementia stage. This broader spread of tau pathology helps explain the higher prevalence of atypical, non-amnestic presentations in younger patients, compared to the more typical amnestic profiles associated with medial temporal tau in late-onset cases–consistent with previous studies (*3, 29*).

The recently updated Alzheimer’s Association criteria for diagnosis and staging introduce an integrated framework pairing biomarker-defined biological stages and clinical stages based on cognitive status (*30*). The extent and spatial distribution of tau PET is presented as the only biomarker validated to date for disease staging. According to the framework, absent tau PET signal characterizes the initial stage and is associated with asymptomatic disease. In contrast, medial temporal lobe–restricted tau defines the early stage, typically linked to transitional or subtle cognitive decline. Tau PET signal in neocortical areas characterizes the intermediate and advanced stages, depending on the degree or spread of binding, and these stages are expected to correspond with a diagnosis of MCI or dementia (Clinical stages 3-6). Our study focused on Aβ-positive cases with MCI or dementia qualifying for EOAD or LOAD diagnosis, who would be expected to have neocortical tau pathology according to the framework. The relatively low tau positivity rates across all neocortical ROIs in LOAD contrast sharply with EOAD’s extensive positivity. This suggests that in LOAD, clinical stage may outpace biological stage, as dementia often associates with pathology confined to early tau areas (i.e., metatemporal ROI), while EOAD patients at the same clinical stage exhibit tau pathology in more advanced regions, such as frontal areas (*31*). However, tau PET positivity should be interpreted cautiously, as the biomarker detects with high accuracy only advanced tau pathology (Braak stage V-VI), often missing early-to-moderate tau patterns (Braak I-IV) (*32*). Despite this limitation, the relatively lower tau positivity rates in LOAD may reflect mixed pathologies. Given the higher co-pathology burden in LOAD (*7*), cognitive decline in older age may be associated with the collective burden of different coexisting pathologies (*33, 34*) rather than exclusively with AD. Our observations support this by showing that cognitive decline correlates closely with tau burden in EOAD (*35*), a relationship that appears less pronounced in LOAD. Intriguingly, the interaction of cognitive scores by age groups to predict tau has not been reported before and supports the idea that with advancing age of symptom onset, clinical outcomes and cognitive decline are not linked solely to tau pathology, a concept that aligns with previous studies in LOAD, where low tau burden was associated with a high prevalence of comorbidities (*36, 37*). Interestingly, this interaction was absent in the medial temporal lobe, the area which typically shows the earliest tau deposition (*38*) and only marginal differences in tau between EOAD and LOAD while adjusting for covariates. One potential explanation for the similarity in baseline and longitudinal medial temporal lobe tau PET binding in EOAD and LOAD, and their similar relationship with demographic and clinical outcomes, is that this region reaches a relative plateau of tau pathology or tau PET binding by the MCI stage in both age groups. Of interest, higher ^18^F-Flortaucipir tau PET binding was associated with increasing age only in the basal ganglia and choroid plexus—areas known to exhibit age-dependent off-target binding (*39*) that is not associated with tau pathology.

Our results underscore age’s differential effect on tau burden in individuals with and without cognitive impairment. In the presence of impairment, EOAD patients exhibit higher tau and faster accumulation than LOAD, suggesting a more aggressive disease course, as discussed above. In contrast, cognitively normal individuals show the opposite pattern, with older participants displaying higher medial temporal lobe tau, consistent with age-related, Aβ-independent increases in tau deposition observed in previous studies (*40*). *APOE*-ε4, a major AD risk factor, is associated with differential tau load and distribution in EOAD and LOAD (*41–43*). In our study, we found no interaction effect of *APOE*-ε4 status and age group on tau load or rate of accumulation. This suggests that although *APOE*-ε4 shapes tau patterns, it does not explain the age-related tau differences in our data. Instead, these differences are likely driven by other factors—such as non-*APOE* genetic factors and distinct neurobiological mechanisms differentiating EOAD and LOAD (*44, 45*).

The faster and more extensive build-up of tau in EOAD compared to LOAD, as shown in our longitudinal analyses, helps explain differences in neurodegeneration and cognitive trajectories between groups. EOAD patients typically experience more widespread and faster atrophy rates (*46*), and more rapid cognitive decline compared to LOAD (*47*), consistent with the accelerated tau accumulation in neocortical areas that we observed in EOAD. Although tau accumulation rates differed between EOAD and LOAD in most cortical regions, the differences were most pronounced in areas beyond the temporoparietal regions (*38, 48*). In the temporoparietal regions, both groups showed tau increases, but with a notable delay in LOAD compared to EOAD. In contrast, only EOAD exhibited significant tau accumulation in frontal and medial occipital regions—areas usually affected during the late stages of the disease (*31*) and predominantly showing atrophy in EOAD (*46*), but not in LOAD at similar clinical stages. These inherent differences in tau dynamics across the age groups are consistent with recent in vitro studies linking distinct tau species to age at symptom onset, with earlier onset associated with more aggressive tau seeding (*16*). Our results support this, indicating that an earlier onset predicts faster tau propagation even within EOAD and LOAD groups. This coupling between baseline extensive pathology and accelerated buildup appears to be a characteristic of tau pathology and aligns with ex vivo studies showing that tau seeding increases as tau spreads to more distant brain areas (*49*), consistent with a prion-like model (*50*).

The differences in tau load, spatial extent, and accumulation rate between EOAD and LOAD have important implications for therapeutic strategies, particularly with the advent of clinically approved monoclonal antibodies targeting Aβ. Evidence from phase 3 clinical trials of the Aβ-targeting antibodies donanemab (in primary analyses) and lecanemab (in post hoc subgroup comparisons), suggests that treatment efficacy is closely tied to baseline tau PET. Greater clinical benefit from Aβ plaque lowering is observed in patients with lower baseline tau, while those with advanced baseline tau experience less benefit (*51, 52*). Our results underscore the need for careful consideration when implementing Aβ-targeting therapies in EOAD. Given the more extensive tau pathology in EOAD, achieving similar benefits as in LOAD may require earlier intervention. The narrower therapeutic window during the symptomatic stage highlights the importance of identifying EOAD at earlier biological stages, when tau pathology is less widespread, and the expected benefit of the antibodies will be greater. Furthermore, the strong correlation between tau and clinical outcomes in EOAD highlights the potential of therapies directly targeting tau, which alone or combined with Aβ-immunotherapies, may slow disease progression in this group (*53*). Despite EOAD patients often being underrepresented in trials due to age limits and the focus on memory loss, which is not always the primary symptom in early onset disease (*3, 28, 29*), these patients may be excellent candidates for testing tau treatments. Their rapid decline (*47*) and strong motivation could facilitate proof-of-concept trials with smaller sample sizes, offering a more efficient path to demonstrating target engagement and cognitive trajectory changes.

From a methodological standpoint, we utilized the CenTauR method for quantifying tau PET binding in template-defined ROIs (*17*). This method standardizes scan processing across laboratories and tracers by employing a common pipeline with five target regions and a reference region. We also applied two proposed linear transformations: one converting SUVR values into CenTauR Z-scores, and another mapping them to the common across tracers CTR unit scale from the Joint Propagation Model approach (*18*)—a method conceptually similar to the Centiloid framework for Aβ PET (*54*). Although tau PET standardization remains an evolving field, with alternative approaches under development, we found that SUVR measures from the CenTauR method were robust and corresponded well to commonly used regions, such as the metatemporal ROI derived from Freesurfer, which is customized to each patient’s anatomy. Additionally, the five different CenTauR ROIs effectively captured the neocortical variability in tau binding patterns observed in the EOAD and LOAD comparison. While further validation is required, and this approach may not suit all purposes, the CenTauR framework shows promise as a standardized measure of tau PET binding.

Strengths of our study include the large sizes of included cohorts, the longitudinal tau PET data, the harmonized procedures adopted from both studies to facilitate comparisons (*55*), and the comprehensive cohort comparisons while adjusting for inherent differences in demographic and clinical characteristics between the EOAD LEADS participants and the LOAD ADNI participants.

Our study also has several limitations. LOAD refers to patients with symptom onset after age 65; however, in ADNI, age of symptom onset is not consistently or reliably reported for all participants. Thus, although we applied a baseline age cutoff of 65 to exclude potential EOAD cases, some misclassification may remain due to limitations in cohort reporting. However, the persistent influence of age across cohorts, evident in cross-sectional and longitudinal tau PET analyses, supports the robustness of our observations. Secondly, despite adjustments for sex and disease stage differences between cohorts, we cannot fully exclude the influence of these factors. Additionally, variances in sample size and follow-up intervals could impact the findings. Finally, the limited racial and ethnic diversity of both cohorts could affect the generalizability of our findings to underrepresented populations.

In conclusion, our study underscores substantial differences in tau pathology between EOAD and LOAD. These differences include higher baseline tau loads, faster tau accumulation over time, and more extensive neuroanatomical involvement in younger patients, suggesting that earlier age of onset in AD is linked to a more aggressive tauopathy. These findings contrast with the relatively smaller differences in Aβ burden across age groups. Our findings contribute to a growing understanding of AD heterogeneity, with age of onset likely serving as a proxy for differences in the biological drivers of disease.

Developing a deeper understanding of the similarities and differences in AD pathophysiology across the age spectrum will advance our ability to identify optimal therapeutic approaches as we enter a new era of disease-modifying therapies in AD.

## Methods

### Participants

All participants were enrolled in either the LEADS (*56*) or the ADNI cohorts. Eligibility for the current analysis required participants to have undergone at least one tau PET scan with ^18^F-Flortaucipir and to possess a complete dataset including Aβ PET and diagnostic data within one year of the baseline tau PET scan for both cohorts. The inclusion criteria were: 1. Patients diagnosed with MCI or dementia due to AD, meeting standard criteria (*57*), and showing a concurrent positive Aβ PET scan, as detailed below, and 2. Cognitively normal individuals, who were included for comparison.

Both ADNI and LEADS were conducted in strict accordance with Good Clinical Practice guidelines and Regulations for the Protection of Human Subjects of Research. Study protocols adhered to the International Conference on Harmonization, the Health Insurance Portability and Accountability Act, and relevant state and federal regulations, receiving approval from local Institutional Review Boards (IRB). Consent was obtained in writing from individuals or their surrogates using IRB-approved forms.

### LEADS

LEADS is a prospective, multisite observational clinical and biomarker study focusing on cognitively impaired participants diagnosed before the age of 65 (NCT03507257) (*55*). It includes participants who meet the 2011 criteria from the National Institute on Aging and the Alzheimer’s Association (NIA-AA) for MCI or mild dementia due to AD (*57*), with a global CDR ≤1, and aged 40–64 years at consent time. Exclusion criteria include known pathogenic mutations in genes associated with autosomal dominant AD (i.e., PSEN1, PSEN2, APP) or familial frontotemporal dementia (Microtubule Associated Protein Tau gene (MAPT), chromosome 9 open reading frame 72 gene (C9ORF72), and Granulin Precursor aka Progranulin (GRN)), which along with AD represents the leading cause of early-onset neurodegenerative dementia (*58*). The criteria for cognitively normal participants include a MMSE score ≥ 24 and a global CDR of 0.

Data for our analysis were sourced from the LEADS repository (https://leads-study.medicine.iu.edu) on March 3, 2025. We identified n=390 Aβ PET positive MCI or dementia patients, who were grouped in an EOAD group. A subset of n=245 participants with EOAD had longitudinal tau PET data with ^18^F-Flortaucipir with an average follow-up interval of 2.06 years. For the subset of EOAD participants who were on Aβ-targeting immunotherapies, biomarker data were used for all time points up until the initiation of treatment. We also identified n=85 and n=9 cognitively normal individuals from the same cohort with negative and positive Aβ PET scans, respectively. These data were used for comparisons of tau PET binding patterns with those observed in the cognitively impaired groups. For a detailed description of the clinical characteristics of the cognitively normal individuals please refer to the Supplementary Table 1.

### ADNI

ADNI is a multicenter study launched in 2003 (NCT02854033) (*59*) and run as a public-private partnership led by Principal Investigator Michael W. Weiner. The primary aim of ADNI is to test whether imaging techniques, biological markers, and clinical and neuropsychological assessment can be combined to measure the progression of mild cognitive impairment (MCI) and early AD. A complete listing of ADNI investigators and participating sites can be found at the end of the article.

Data were obtained from the ADNI repository (https://adni.loni.usc.edu) on February 28, 2025. We identified 211 Aβ PET positive patients diagnosed with MCI or dementia, after excluding 24 cases due to their age being under 65 at the baseline tau PET scan. These patients were categorized into the LOAD group. Notably, this grouping likely includes some individuals, inadvertently, whose symptom-onset occurred before age 65. This categorization was necessary due to the cohort’s lack of a consistent and reliable methodology for precisely determining the age at symptom onset. A subset of n=99 participants with LOAD had longitudinal tau PET data with ^18^F-Flortaucipir with an average follow-up interval of 2.80 years. Additionally, we identified n=328 and n=125 cognitively normal individuals from the same cohort with negative and positive Aβ PET scans, respectively. These data were used for comparisons of tau PET binding patterns with those observed in the cognitively impaired groups. For a detailed description of the clinical characteristics of the cognitively normal individuals please refer to the Supplementary Table 1.

### Cognitive measures

Global cognition was assessed using the MMSE, the CDR-SB, and global CDR scores, all measured within a year of the baseline tau PET scans for both cohorts.

### PET acquisition

Baseline and longitudinal tau PET scans, along with baseline Aβ PET scans, were included in our analyses. The protocols for PET scan acquisition are detailed in prior publications (*37, 56*). Briefly, Aβ PET scans were acquired using ^18^F-florbetaben at 90 to 110 minutes post-injection for LEADS, and ^18^F-florbetapir at 50 to 70 minutes or ^18^F-florbetaben at 90 to 110 minutes post-injection for ADNI. Tau PET scans for both cohorts were performed with ^18^F-flortaucipir at 75 to 105 minutes post-injection. Image acquisition, preprocessing, and quality control are harmonized across ADNI and LEADS.

### PET image analysis

Each PET scan was realigned, averaged, and smoothed to achieve a uniform resolution of 6 mm full width at half maximum (FWHM). The PET images were independently processed following the Centiloid and CenTauR protocols (*17, 54*) for Aβ and ^18^F-flortaucipir tau scans, respectively. Briefly, PET scans were co-registered to the closest in time acquired T1 MRI scan for each individual and time point. The transformation matrix derived from the SPM8 unified segmentation of these T1 MRI images was used to normalize the PET images into the MNI-152 space.

For Aβ PET scans, the template-based Centiloid global cortical and whole cerebellum masks served as target and reference ROI (*54*), respectively. Standardized uptake value ratios (SUVRs) were converted to Centiloids (CL) using established linear equations (*54, 60, 61*), setting a threshold of 24.4 CL to define amyloid positivity for both cohorts (*62*).

For tau PET scans, the CenTauR protocol employed template-based mesial temporal, meta-temporal, temporoparietal, frontal, and universal masks as target ROIs, with the inferior cerebellum mask as the reference (*17*) (Supplementary Fig. 9). SUVRs were scaled to CenTauR Z scores and CenTauR (CTR) units from the Joint Propagation Model approach based on previously published linear equations (*17, 18*)—two methods harmonizing tau PET quantification across tracers. Although our study used only one tau PET tracer (i.e., ^18^F-Flortaucipir), we reported both harmonized measures alongside raw SUVRs to enhance comparability with future observational studies and clinical trials. An exploratory threshold of 2 CenTauR Z scores was set as a potential threshold for tau positivity across ROIs. Comparisons of quantification using the CenTauR meta-temporal target and inferior cerebellum reference masks were made against standard Freesurfer-based quantification in the respective ROIs for the LEADS participants and are presented in Supplementary Fig. 10 (*56*), along with a similar comparison for the Aβ PET processing method.

The normalized tau PET scans were further smoothed using a Gaussian kernel of 4 mm FWHM to prepare for voxel-wise statistical evaluations.

### Statistical analyses – cross-sectional

Demographic and clinical characteristics across age groups were analyzed using independent two-sample t-tests for continuous variables and chi-square tests for categorical variables. We assessed ROI-based differences in tau load between EOAD and LOAD patients at the MCI and dementia stages with pairwise t-tests. Cross-sectional voxel-wise differences in ^18^F-flortaucipir binding were examined using linear regression models, controlling for sex, MMSE score, educational attainment, and Aβ PET load. The relationship between baseline age and ^18^F-flortaucipir binding load was analyzed using separate linear regressions in EOAD and LOAD for ROI-based analyses and in the whole sample for voxel-wise analyses while adjusting for the above-mentioned covariates. Linear regressions at the ROI– and voxel-based levels assessed the interaction between Aβ PET load or cognitive performance (i.e., MMSE, CDR-SB) and age group (EOAD vs. LOAD) in predicting ^18^F-flortaucipir binding, while adjusting for sex, educational attainment, and baseline MMSE or baseline Aβ PET, respectively. Post hoc simple slopes analyses were applied to the regression models assessing the interaction term, for calculating the individual slopes for EOAD and LOAD. For all voxel-wise analyses, corrections for multiple comparisons were performed using the Random Field Theory Family-Wise Error (FWE) correction method (p < 0.05) in the SPM12 toolbox.

### Statistical analyses – longitudinal

We employed linear mixed-effects models to assess the interaction between age group (EOAD and LOAD) and time from baseline on longitudinal ^18^F-flortaucipir binding load changes. These models accounted for participant-specific variations over time through random intercepts and slopes and were adjusted for sex, educational attainment, baseline MMSE, and Aβ PET load. Post-hoc simple slope analyses were conducted to examine the annual rates of change in ^18^F-flortaucipir binding in the EOAD and LOAD groups and across the subgroups of patients at the clinical stages of MCI and dementia. Separate linear mixed-effects models for EOAD and LOAD estimated the average annual rate of binding change for each participant, and the association of those as a function of baseline age was assessed with Pearson’s correlation. Similar linear mixed-effects models were applied in the CenTauR ROIs and in a voxel-wise manner, with corrections for multiple comparisons using Random Field Theory FWE correction method (p < 0.05), as implemented in the Voxelstats toolbox (*20*).

We used the Sampled Iterative Local Approximation (SILA) algorithm (*21*) for modeling separately the ^18^F-flortaucipir tau PET trajectories of EOAD and LOAD. SILA employs discrete sampling of longitudinal ^18^F-Flortaucipir tau PET binding versus age data to model the relationship between binding rates and levels. This algorithm uses numerical robust smoothing techniques and Euler’s method for numerical integration, generating non-parametric tau load versus tau positivity duration (i.e., estimated tau years) curves, with time zero corresponding to the tau positivity threshold of 2 CenTauR Z scores in the MetaTemporal ROI.

## List of Supplementary Materials

**Supplementary Table 1**. Clinical characteristics of the cognitively normal (CN) groups from the LEADS and ADNI cohorts stratified by Aβ PET status.

**Supplementary Table 2**. Annual rates of change in ^18^F-Flortaucipir tau PET SUVR binding deriving from the linear mixed-effects models analyses in the whole sample (i.e. EOAD and LOAD), and separately in cases at the MCI and dementia stages at baseline.

**Supplementary Fig. 1**. Group comparisons of baseline ^18^F-Flortaucipir binding across EOAD and LOAD patient groups in the temporo-parietal CenTauR ROI. Formal statistical tests were restricted to EOAD and LOAD groups, with cognitively normal (CN) participants plotted for illustration purposes. The percentages above each boxplot indicate the proportion of individuals exceeding the +2 CenTauR Z-score threshold for binding load, based on Villemagne et al. The data are also presented on the CenTauR (CTR) scale from the Joint Propagation Model approach, following Leuzy et al.

**Supplementary Fig. 2**. Average baseline^18^F-Flortaucipir tau PET binding images in the groups of patients with EOAD and LOAD stratified by global CDR scores.

**Supplementary Fig. 3**. Voxel-wise group comparison illustrating regions where patients with LOAD show greater baseline ^18^F-Flortaucipir tau PET binding compared to those with EOAD. Patients missing baseline MMSE scores were excluded. These voxel-wise analyses were adjusted for baseline Centiloid, MMSE, sex, years of education, and corrected for multiple comparisons using FWE rate at the peak level, as implemented in SPM12 software.

**Supplementary Fig. 4**. Scatterplots showing the association between age and baseline ^18^F-Flortaucipir tau PET binding at the clinical stages of MCI and dementia, across two age groups (i.e., EOAD and LOAD), in the CenTauR ROIs for the mesial temporal (A) and universal masks (B). Linear models were adjusted for baseline Centiloid, MMSE, sex, and years of education. Patients missing baseline MMSE scores were excluded. The linear coefficients for EOAD and LOAD are shown in orange and blue, respectively.

**Supplementary Fig. 5**. Voxel-wise associations between age and baseline ^18^F-Flortaucipir tau PET binding load across the entire sample. Patients missing baseline MMSE scores were excluded. These voxel-wise analyses were adjusted for baseline Centiloid, MMSE, sex, and years of education, and corrected for multiple comparisons using the FWE rate at the peak level, as implemented in SPM12 software.

**Supplementary Fig. 6.** Longitudinal modeling of ^18^F-Flortaucipir tau PET binding in EOAD and LOAD, illustrated by trajectory plots for individual patients (A), scatterplots showing age at baseline relative to the estimated annual rate of change in tau PET binding from the linear mixed-effects models (B). The linear mixed-effects models (B) were adjusted for baseline MMSE score, Centiloid, sex, and years of education, with random intercepts and slopes at the individual patient level. Patients missing baseline MMSE scores were excluded from the statistical modeling. The estimated annual rates of change in ^18^F-Flortaucipir tau PET binding were derived from the sum of the fixed and random coefficients for each individual. The Pearson correlation coefficients for EOAD and LOAD are shown in orange and blue, respectively.

**Supplementary Fig. 7.** Longitudinal changes in ^18^F-Flortaucipir tau PET binding over the follow-up interval in patients with EOAD and LOAD, stratified by *APOE*-ε4 status. The linear mixed-effects were adjusted for baseline MMSE score, Centiloid, sex, and years of education, with random intercepts and slopes at the individual patient level. Patients missing baseline MMSE scores were excluded from the statistical modeling. The analyses were corrected for multiple comparisons using the FWE rate at the peak level, as implemented in Voxelstats software.

**Supplementary Fig. 8.** Longitudinal modeling of ^18^F-Flortaucipir tau PET binding in EOAD and LOAD, when modeling group trajectories as a function of tau duration from the CenTauR threshold of +2 Z-scores using the SILA pipeline. The SILA models were anchored to the +2 CenTauR Z-score threshold, which was used to define ^18^F-Flortaucipir tau PET binding positivity, and the trajectories are modeled relative to time in years from crossing the threshold.

**Supplementary Fig. 9**. Standard template-based masks for Centiloid Aβ PET and CenTauR ^18^F-Flortaucipir PET quantification. Notably, several CenTauR masks show spatial overlap. The Centiloid and CenTauR methodologies also differ in their reference region masks for the cerebellum: Centiloid uses the whole cerebellum, while CenTauR uses the inferior cerebellum.

**Supplementary Fig. 10**. Association between Freesurfer-based and standard template-based masks for quantifying ^18^F-Flortaucipir PET and ^18^F-Florbetaben PET binding in a subset of LEADS participants (n=460 out of 477), following the CenTauR and Centiloid pipelines, respectively.

## Data Availability

The anonymized data that support the findings of this study are available on request from the LEADS data core (https://leads-study.medicine.iu.edu/researchers/leads-data-request-application/) and the ADNI website (https://adni.loni.usc.edu/).

## Acknowledgements

We thank participants and families for their trust and commitment to research. Avid Radiopharmaceuticals enabled the use of Flortaucipir but did not provide direct funding and were not involved in data analysis or interpretation. Life Molecular Imaging enabled the use of Florbetaben but did not provide direct funding and were not involved in data analysis or interpretation. JL received a grant from the Fondation Recherche Alzheimer for his fellowship at UCSF.

## Author contributions

Conceptualization: LGA, BCD, GDR., MCC, RLJ, and KC

Design: LGA, BCD, GDR., MCC, RLJ, and KC

Acquisition of the of the data: All authors

Writing – original draft: KC, RLJ, and GDR

Writing – review & editing: All authors

## Supplementary Data

**Supplementary Table 1.** Clinical characteristics of the cognitively normal (CN) groups from the LEADS and ADNI cohorts stratified by Aβ PET status.

| Characteristic | CN (A-) |  |  | CN (A+) |  |  |
| --- | --- | --- | --- | --- | --- | --- |
|  | LEADS,<br>N = 85<sup>1</sup> | ADNI,<br>N = 328<sup>1</sup> | p-value<sup>2</sup> | LEADS,<br>N = 9<sup>1</sup> | ADNI,<br>N = 125<sup>1</sup> | p-value<sup>2</sup> |
| **Age (years)** | 56.80 ± 5.79 | 73.27 ± 6.10 | **<0.001** | 59.94 ± 5.82 | 75.57 ± 6.90 | **<0.001** |
| **Sex** |  |  | 0.300 |  |  | 0.600 |
| Male | 30 (35.29%) | 140 (42.68%) |  | 5 (55.56%) | 50 (40.00%) |  |
| Female | 55 (64.71%) | 188 (57.32%) |  | 4 (44.44%) | 75 (60.00%) |  |
| **Education in years** | 16.69 ± 1.92 | 16.93 ± 2.27 | 0.300 | 16.33 ± 3.67 | 16.59 ± 2.23 | 0.800 |
| **APOE-ε4 allele** |  |  | **0.014** |  |  | **<0.001** |
| Carrier (≥ 1 allele) | 32 (38.09%) | 69 (23.07%) | | 7 (77.78%) | 62 (53.45%) | |
| Non carrier | 52 (61.90%) | 230 (76.92%) |  | 2 (22.22%) | 54 (46.55%) |  |
| Missing | 1 | 29 |  | 0 | 9 |  |
| **Race/Ethnicity** |  |  | 0.500 |  |  | 0.200 |
| White, not Hispanic or Latino | 54 (64.29%) | 233 (73.97%) |  | 6 (66.67%) | 101 (86.32%) |  |
| White, Hispanic or Latino | 7 (8.33%) | 17 (5.40%) |  | 0 (0.00%) | 3 (2.56%) |  |
| Black or African American | 14 (16.67%) | 43 (13.65%) |  | 3 (33.33%) | 8 (6.84%) |  |
| Asian | 6 (7.14%) | 13 (4.13%) |  | 0 (0.00%) | 2 (1.71%) |  |
| Other | 0 (0.00%) | 2 (0.63%) |  | 0 (0.00%) | 1 (0.85%) |  |
| More than one | 3 (3.57%) | 7 (2.22%) |  | 0 (0.00%) | 2 (1.71%) |  |
| Missing | 1 | 13 |  | 0 | 8 |  |
| **Centiloids** | 1.82 ± 6.50 | 0.59 ± 10.95 | 0.200 | 43.80 ± 13.66 | 63.20 ± 30.74 | **0.002** |
| **CDR-SB** | 0.01 ± 0.08 | 0.06 ± 0.22 | **<0.001** | 0.00 ± 0.00 | 0.07 ± 0.20 | **<0.001** |
| Missing | 0 | 2 |  | 0 | 0 |  |
| **CDR Global** |  |  | 0.200 |  |  | >0.900 |
| 0 | 85 (100.00%) | 315 (96.63%) |  | 9 (100.00%) | 119 (95.20%) |  |
| 0.5 | 0 (0.00%) | 11 (3.37%) |  | 0 (0.00%) | 6 (4.80%) |  |
| Missing | 0 | 2 |  | 0 | 0 |  |
| **MMSE** | 29.20 ± 0.94 | 29.12 ± 1.13 | 0.500 | 29.00 ± 0.87 | 28.89 ± 1.32 | 0.700 |
| Missing | 1 | 2 |  | 0 | 0 |  |

<sup>1</sup>Mean ± SD; n (%); <sup>2</sup>Welch Two Sample t-test; Pearson's Chi-squared test. CDR = Clinical Dementia Rating; MMSE = Mini Mental State Examination

**Supplementary Table 2.** Annual rates of change in ^18^F-Flortaucipir tau PET SUVR binding deriving from the linear mixed-effects models analyses in the whole sample (i.e. EOAD and LOAD), and separately in cases at the MCI and dementia stages at baseline.

|  | Mesial temporal |  | Meta- temporal |  | TemporoParietal |  | Frontal |  | Universal |  |
| --- | --- | --- | --- | --- | --- | --- | --- | --- | --- | --- |
|  | Slope ± SE<br>(SUVR/yea<br>r) | p | Slope ± SE<br>(SUVR/yea<br>r) | p | Slope ± SE<br>(SUVR/yea<br>r) | p | Slope ± SE<br>(SUVR/yea<br>r) | p | Slope ± SE<br>(SUVR/yea<br>r) | p |
| All |  |  |  |  |  |  |  |  |  |  |
| EOAD (n=243) | 0.06±0.00 | <0.001 | 0.07±0.01 | <0.001 | 0.06±0.01 | <0.001 | 0.10±0.01 | <0.001 | 0.07±0.01 | <0.001 |
| LOAD (n=99) | 0.02±0.01 | 0.008 | 0.04±0.01 | <0.001 | 0.03±0.01 | 0.004 | 0.02±0.01 | 0.111 | 0.03±0.01 | 0.003 |
| Interaction (Δ) | 0.05±0.01 | <0.001 | 0.04±0.01 | 0.001 | 0.03±0.01 | 0.013 | 0.08±0.01 | <0.001 | 0.04±0.01 | 0.001 |
| MCI stage at baseline |  |  |  |  |  |  |  |  |  |  |
| EOAD (n=66) | 0.05 ± 0.01 | <0.001 | 0.08±0.01 | <0.001 | 0.07±0.01 | <0.001 | 0.08±0.01 | <0.001 | 0.07±0.01 | <0.001 |
| LOAD (n=64) | 0.02 ± 0.01 | <0.001 | 0.03±0.01 | <0.001 | 0.02±0.01 | <0.001 | 0.01±0.01 | 0.519 | 0.02±0.01 | <0.001 |
| Interaction (Δ) | 0.03 ± 0.01 | <0.001 | 0.05±0.01 | <0.001 | 0.05±0.01 | <0.001 | 0.07±0.01 | <0.001 | 0.05±0.01 | <0.001 |
| Dementia stage at baseline |  |  |  |  |  |  |  |  |  |  |
| EOAD (n=177) | 0.06±0.01 | <0.001 | 0.07±0.01 | <0.001 | 0.05±0.01 | <0.001 | 0.10±0.01 | <0.001 | 0.06±0.01 | <0.001 |
| LOAD (n=35) | 0.00±0.01 | 0.738 | 0.02±0.01 | 0.211 | 0.01±0.02 | 0.403 | 0.01±0.02 | 0.527 | 0.01±0.02 | 0.436 |
| Interaction (Δ) | 0.06±0.01 | <0.001 | 0.05±0.02 | 0.004 | 0.04±0.02 | 0.053 | 0.08±0.02 | <0.001 | 0.05±0.02 | 0.010 |

**Supplementary Fig. 1.**
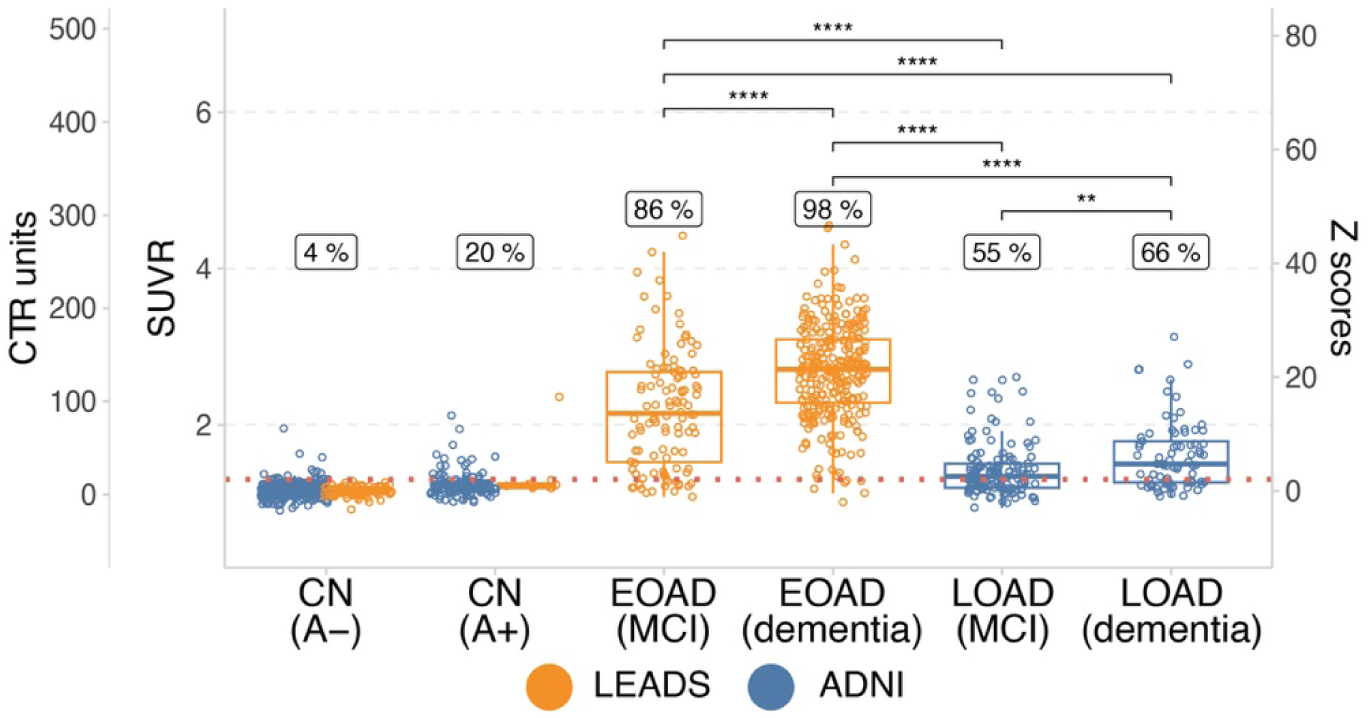
Group comparisons of baseline ^18^F-Flortaucipir binding across EOAD and LOAD patient groups in the temporo-parietal CenTauR ROI. Formal statistical tests were restricted to EOAD and LOAD groups, with cognitively normal (CN) participants plotted for illustration purposes. The percentages above each boxplot indicate the proportion of individuals exceeding the +2 CenTauR Z-score threshold for binding load, based on Villemagne et al. The data are also presented on the CenTauR (CTR) scale from the Joint Propagation Model approach, following Leuzy et al.

**Supplementary Fig. 2.**
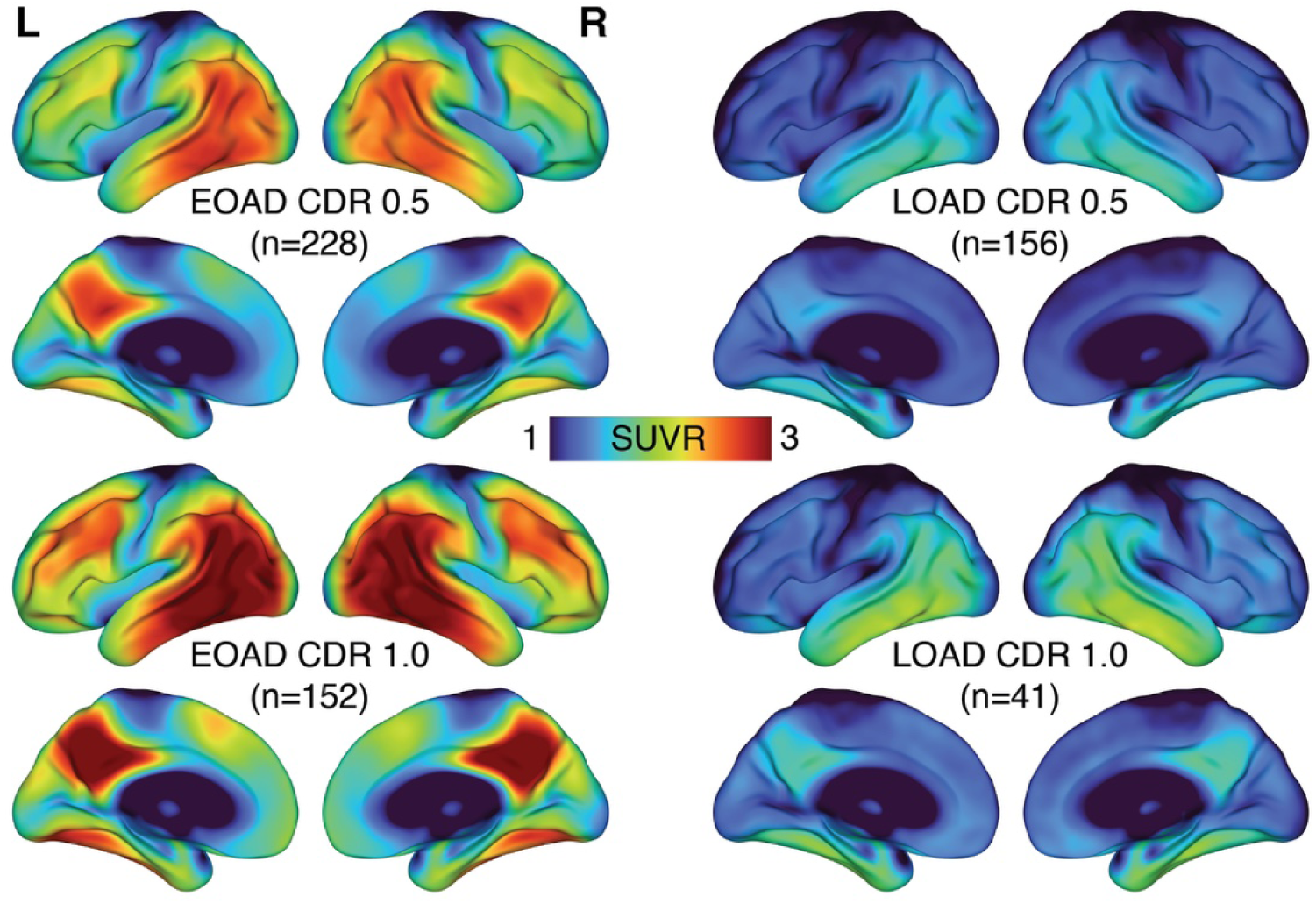
Average baseline^18^F-Flortaucipir tau PET binding images in the groups of patients with EOAD and LOAD stratified by global CDR scores.

**Supplementary Fig. 3.**
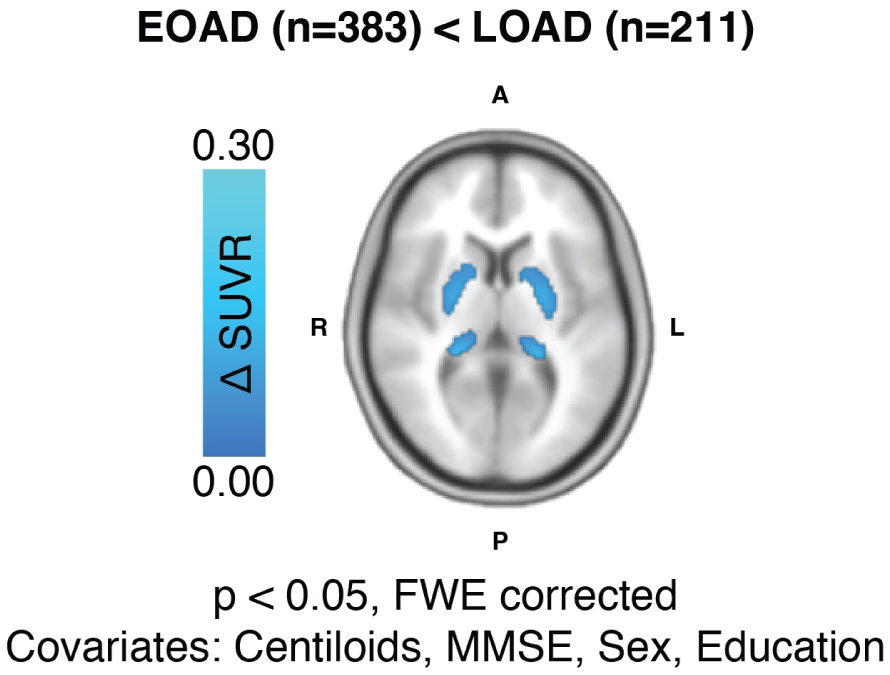
Voxel-wise group comparison illustrating regions where patients with LOAD show greater baseline ^18^F-Flortaucipir tau PET binding compared to those with EOAD. Patients missing baseline MMSE scores were excluded. These voxel-wise analyses were adjusted for baseline Centiloid, MMSE, sex, years of education, and corrected for multiple comparisons using FWE rate at the peak level, as implemented in SPM12 software.

**Supplementary Fig. 4.**
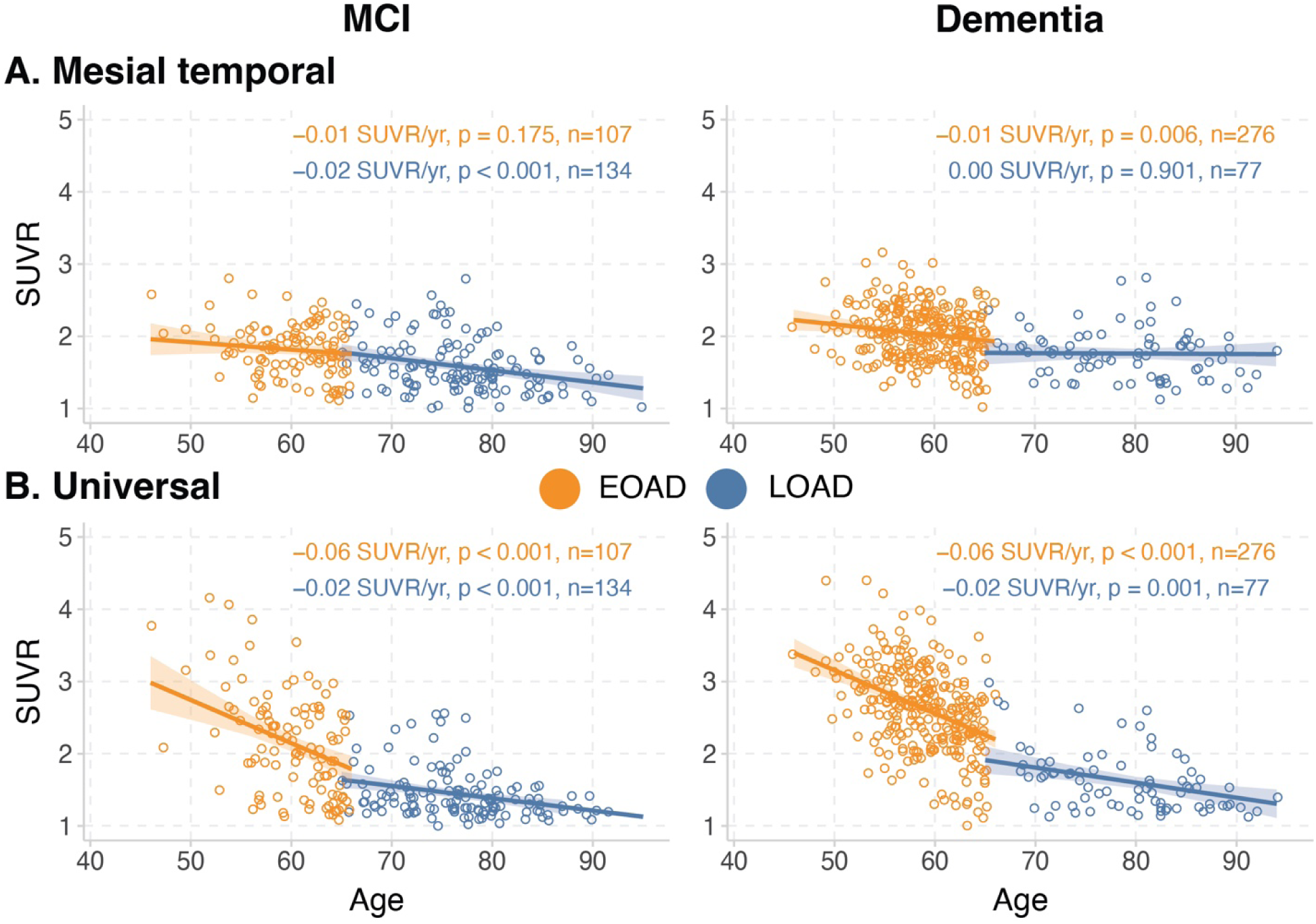
Scatterplots showing the association between age and baseline ^18^F-Flortaucipir tau PET binding at the clinical stages of MCI and dementia, across two age groups (i.e., EOAD and LOAD), in the CenTauR ROIs for the mesial temporal. (A) and universal masks (B). Linear models were adjusted for baseline Centiloid, MMSE, sex, and years of education. Patients missing baseline MMSE scores were excluded. The linear coefficients for EOAD and LOAD are shown in orange and blue, respectively.

**Supplementary Fig. 5.**
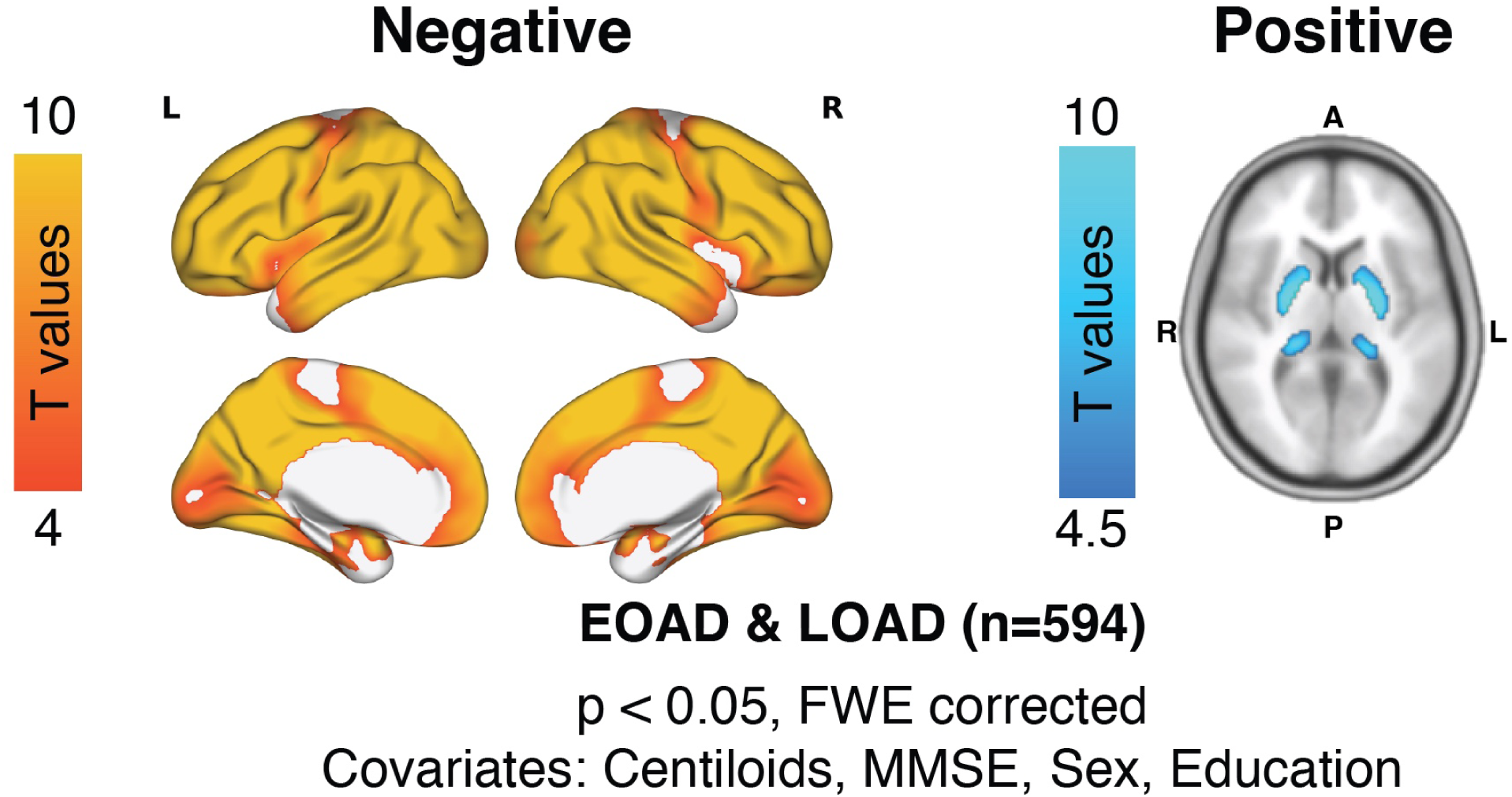
Voxel-wise associations between age and baseline ^18^F-Flortaucipir tau PET binding load across the entire sample. Patients missing baseline MMSE scores were excluded. These voxel-wise analyses were adjusted for baseline Centiloid, MMSE, sex, and years of education, and corrected for multiple comparisons using the FWE rate at the peak level, as implemented in SPM12 software.

**Supplementary Fig. 6.**
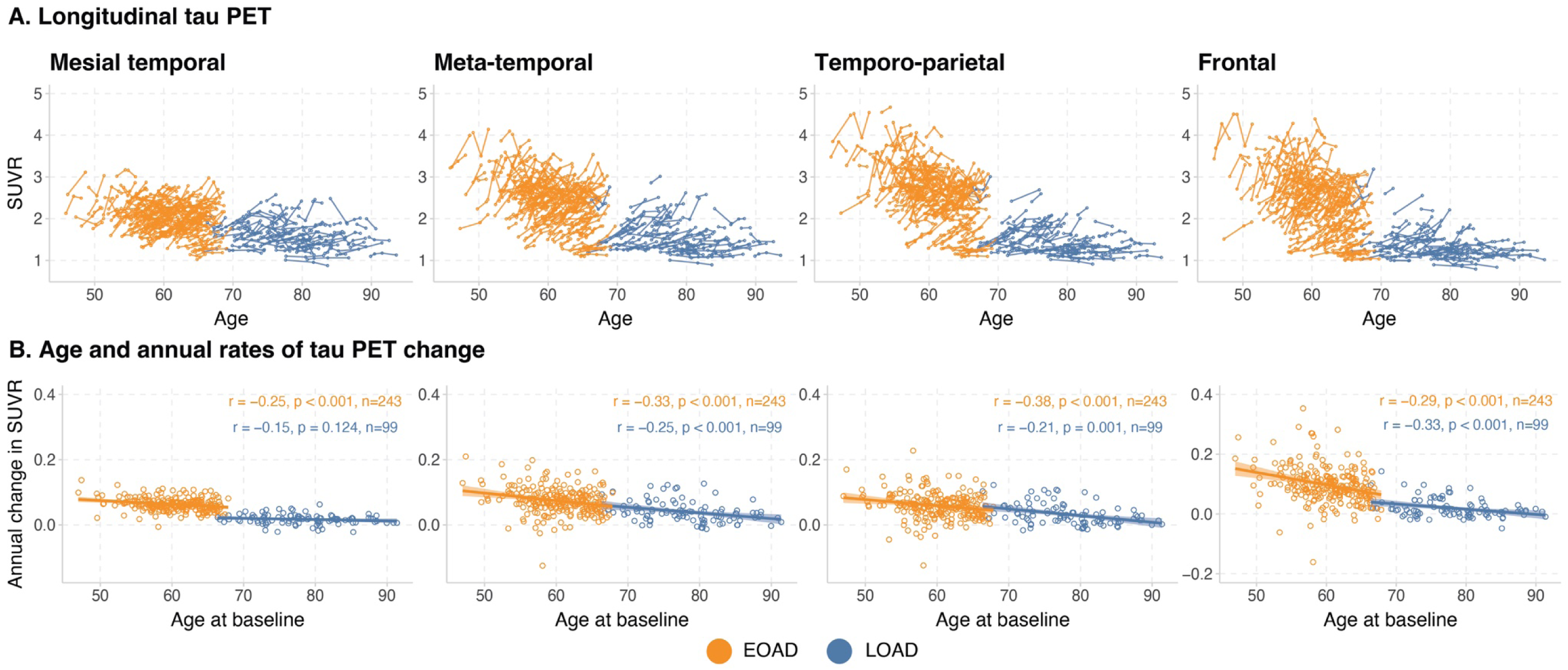
Longitudinal modeling of ^18^F-Flortaucipir tau PET binding in EOAD and LOAD, illustrated by trajectory plots for individual patients. (A), scatterplots showing age at baseline relative to the estimated annual rate of change in tau PET binding from the linear mixed-effects models (B). The linear mixed-effects models (B) were adjusted for baseline MMSE score, Centiloid, sex, and years of education, with random intercepts and slopes at the individual patient level. Patients missing baseline MMSE scores were excluded from the statistical modeling. The estimated annual rates of change in ^18^F-Flortaucipir tau PET binding were derived from the sum of the fixed and random coefficients for each individual. The Pearson correlation coefficients for EOAD and LOAD are shown in orange and blue, respectively.

**Supplementary Fig. 7.**
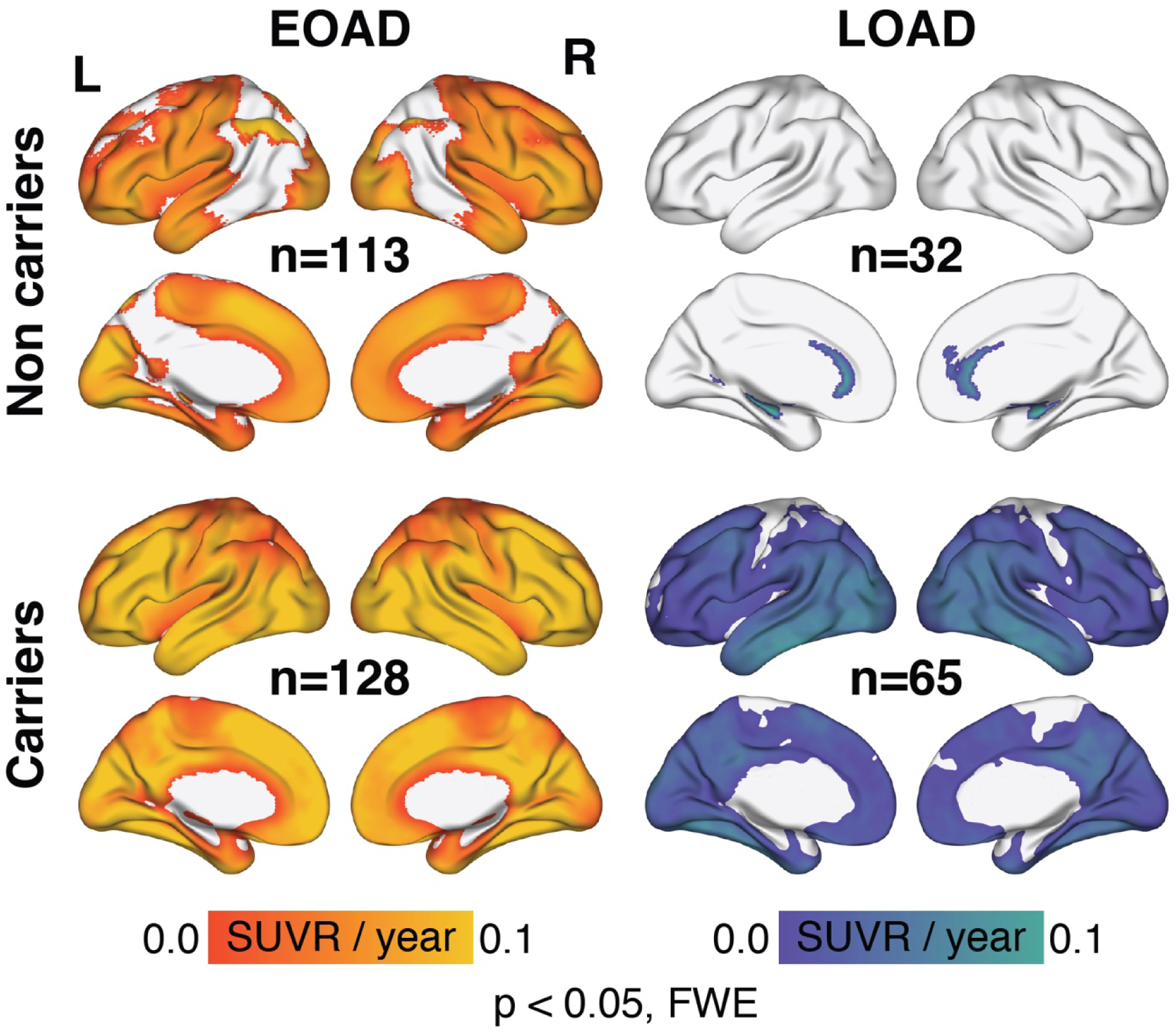
Longitudinal changes in ^18^F-Flortaucipir tau PET binding over the follow-up interval in patients with EOAD and LOAD, stratified by *APOE*-ε4 status. The linear mixed-effects were adjusted for baseline MMSE score, Centiloid, sex, and years of education, with random intercepts and slopes at the individual patient level. Patients missing baseline MMSE scores were excluded from the statistical modeling. The analyses were corrected for multiple comparisons using the FWE rate at the peak level, as implemented in Voxelstats software.

**Supplementary Fig. 8.**
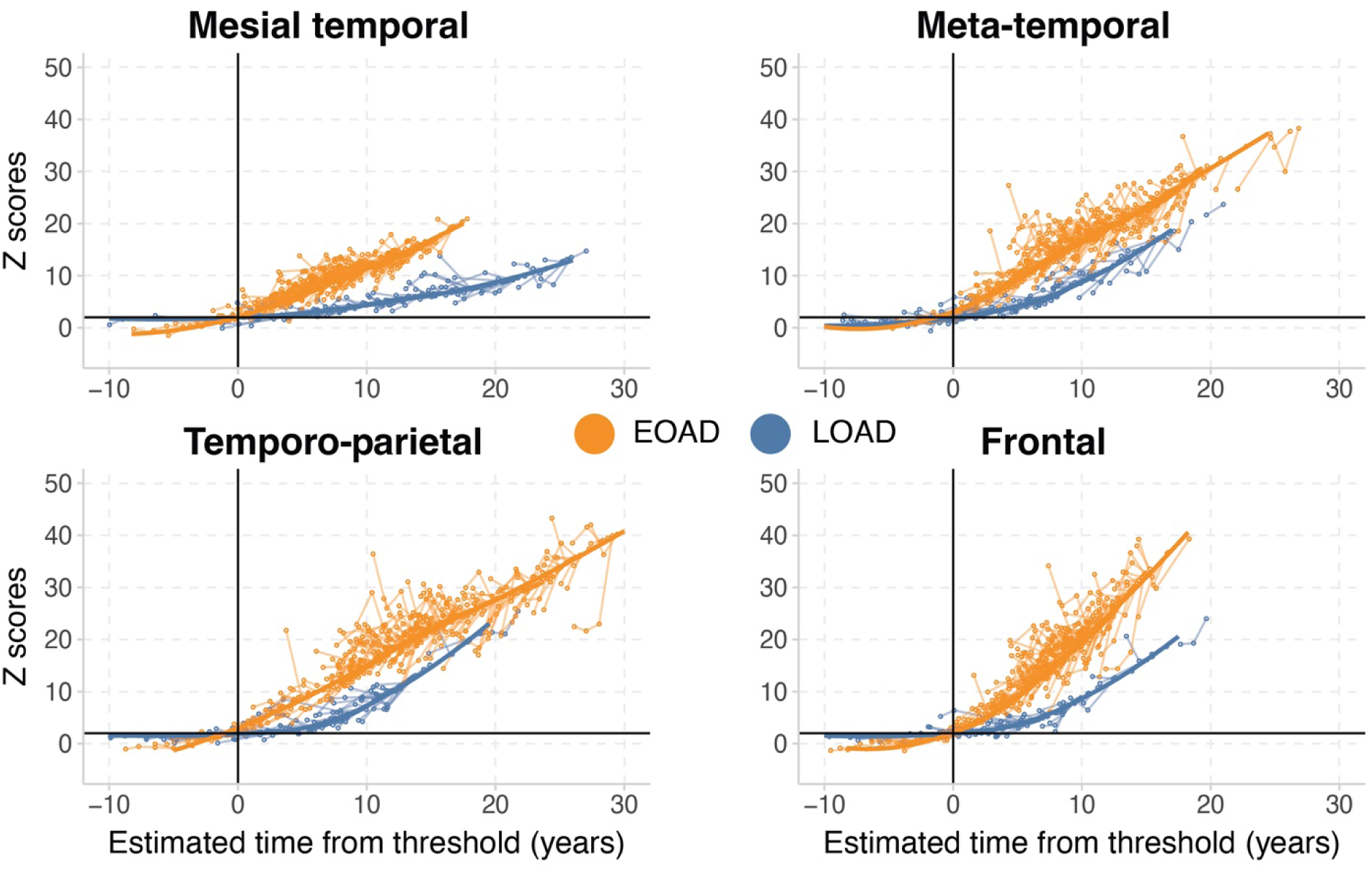
Longitudinal modeling of ^18^F-Flortaucipir tau PET binding in EOAD and LOAD, when modeling group trajectories as a function of tau duration from the CenTauR threshold of +2 Z-scores using the SILA pipeline. The SILA models were anchored to the +2 CenTauR Z-score threshold, which was used to define ^18^F-Flortaucipir tau PET binding positivity, and the trajectories are modeled relative to time in years from crossing the threshold.

**Supplementary Fig. 9.**
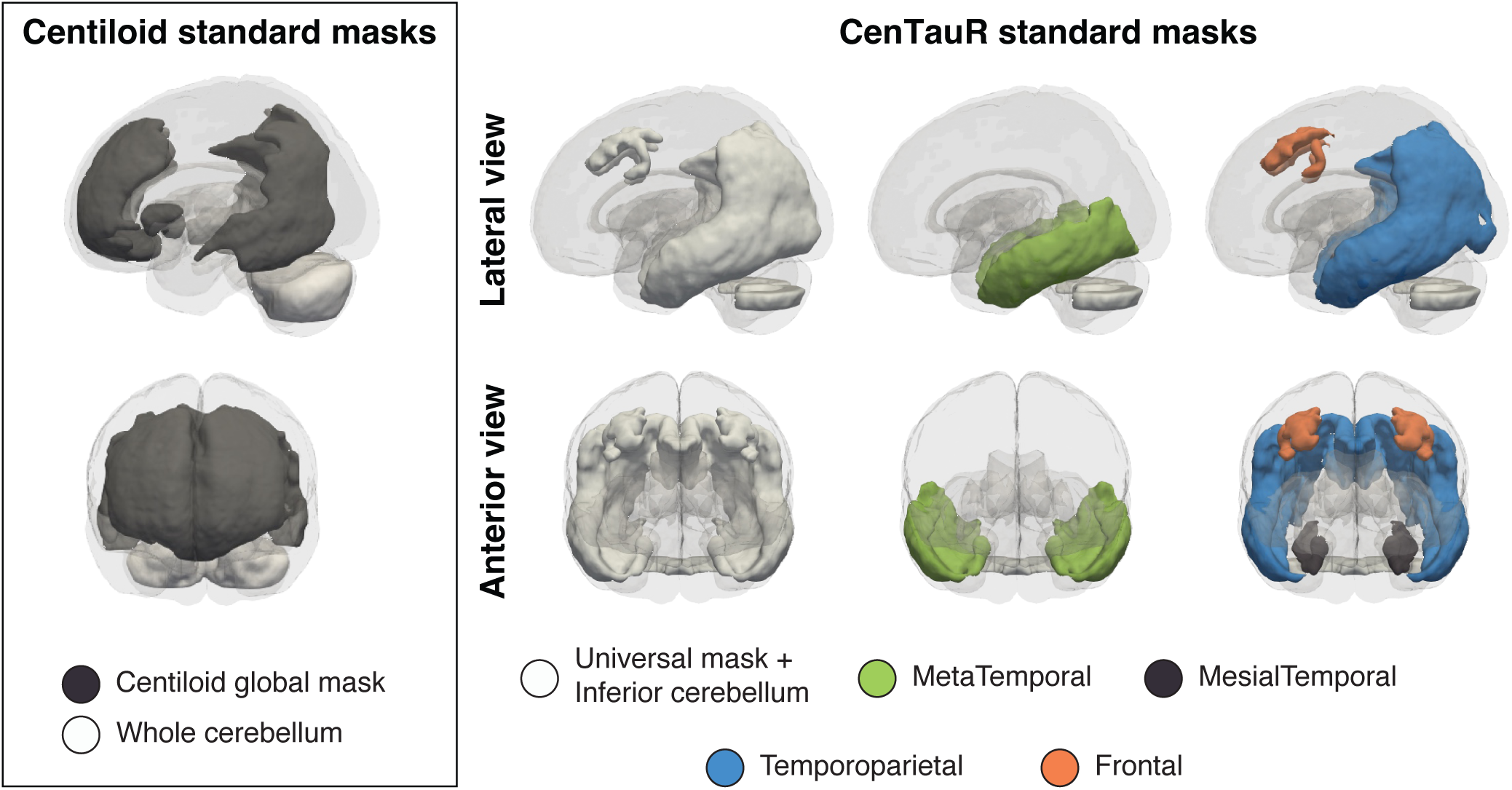
Standard template-based masks for Centiloid Aβ PET and CenTauR ^18^F-Flortaucipir PET quantification. Notably, several CenTauR masks show spatial overlap. The Centiloid and CenTauR methodologies also differ in their reference region masks for the cerebellum: Centiloid uses the whole cerebellum, while CenTauR uses the inferior cerebellum.

**Supplementary Fig. 10.**
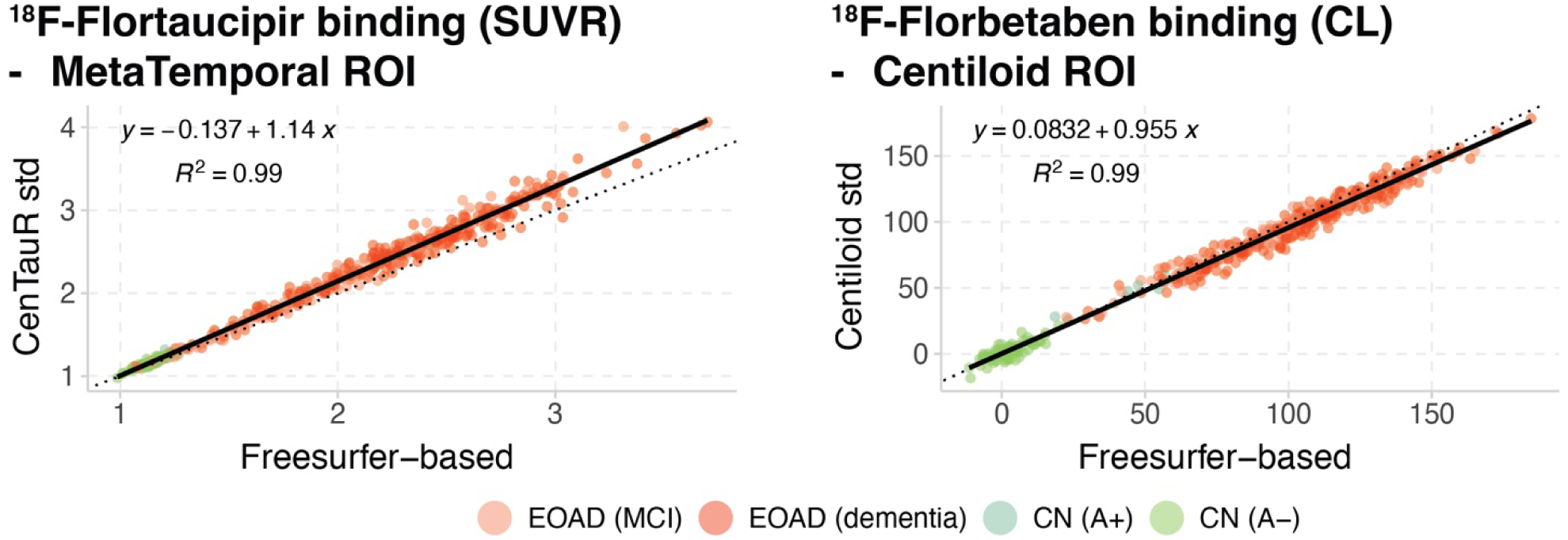
Association between Freesurfer-based and standard template-based masks for quantifying ^18^F-Flortaucipir PET and ^18^F-Florbetaben PET binding in a subset of LEADS participants (n=460 out of 477), following the CenTauR and Centiloid pipelines, respectively.

